# Evidence of survival bias in the association between *APOE-Є4* and age of ischemic stroke onset

**DOI:** 10.1101/2023.12.01.23294385

**Authors:** Joanna von Berg, Patrick F. McArdle, Paavo Häppölä, Jeffrey Haessler, Charles Kooperberg, Robin Lemmens, Alessandro Pezzini, Vincent Thijs, SiGN, FinnGen, Women’s Health Initiative, Sara L. Pulit, Steven J. Kittner, Braxton D. Mitchell, Jeroen de Ridder, Sander W. van der Laan

## Abstract

Large genome-wide association studies (GWAS) employing case-control study designs have now identified tens of loci associated with ischemic stroke (IS). As a complement to these studies, we performed GWAS in a case-only design to identify loci influencing age at onset (AAO) of ischemic stroke. Analyses were conducted in a Discovery cohort of 10,857 ischemic stroke cases using a linear regression framework. We meta-analyzed all SNPs with p-value < 1×10^−5^ in a sex-combined or sex-stratified analysis using summary data from two additional replication cohorts. In the women-only meta-analysis, we detected significant evidence for association of AAO with rs429358, an exonic variant in *APOE* that encodes for the APOE-є4 allele. Each copy of the rs429358:T>C allele was associated with a 1.29 years earlier stroke AOO (meta p-value = 2.48×10^−11^).

This *APOE* variant has previously been associated with increased mortality and ischemic stroke AAO. We hypothesized that the association with AAO may reflect a survival bias attributable to an age-related decline in mortality among APOE-є4 carriers and have no association to stroke AAO per se. Using a simulation study, we found that a variant associated with overall mortality might indeed be detected with an AAO analysis. A variant with a two-fold increase on mortality risk would lead to an observed effect of AAO that is comparable to what we found. In conclusion, we detected a robust association of the *APOE* locus with stroke AAO and provided simulations to suggest that this association may be unrelated to ischemic stroke per se but related to a general survival bias.

## Introduction

Genetic association analysis and, in particular, genome-wide genetic association studies (GWAS), have become standard tools for identifying genetic contributions to complex diseases, including ischemic stroke. While such studies are typically framed as case-control study designs, case-only designs have also been used, for example, for the purpose of identifying variants that associate with age of disease onset, *i*.*e*., timing of disease. Such associations may reflect variants that predispose to earlier forms of disease or modify the effects of other disease-predisposing variants. Case-only age at onset association analyses have been carried out on multiple traits, including Alzheimer’s disease^1^, Parkinson’s Disease^2^, as well as stroke^3^. Case-control approaches that condition on clinical covariates, such as age at onset, have also been used to boost power to detect risk loci for complex diseases^4,5^.

To identify variants associated with ischemic stroke age at onset (AAO), we performed a two-stage meta-analysis of GWAS for AAO in 10,857 stroke cases from SiGN^6^, followed by a replication of all associated SNPs (p-value < 5×10^−5^) in two independent studies, FinnGen Data Freeze 5^7^ and the Women’s Health Initiative (WHI)^8^. We performed sex-combined and sex-stratified analyses. From these analyses, we identified a variant in the *APOE* locus, encoding the ApoE-є4 allele, that was significantly associated with earlier AAO in women (rs429358:T>C, meta p-value = 2.48×10^−11^, beta = 1.29 ± 0.38 years), but not in men. This SNP has previously been associated with stroke AAO in a prior candidate gene study^3^, yet is not associated with risk of ischemic stroke (OR=1.0, 0.97-1.03 95% confidence interval, p = 0.97, n_cases_=33,936 vs. n_controls_=391,114 in European populations^9^). Thus, we hypothesized that the association with earlier stroke AAO may reflect an overall association of this variant with earlier death^10^. To test this hypothesis, we performed a simulation study in which we simulated loci that are associated with overall mortality unrelated to stroke to characterize parameters that would lead to age-related differences in allele frequencies that could be misinterpreted as being related to age of disease onset.

## Results

Summary characteristics of the Discovery (SiGN) and follow-up replication cohorts (FinnGen Data Freeze 5 and WHI) are shown in **Table 1**. Mean age of stroke onset was 67.6, 66.7, and 76.6 yrs, in SIGN, FinnGen, and WHI (the latter includes only women), respectively.

**Table 1:** Characteristics of the study populations. Percentages have been calculated on the subset of individuals with no missing values for the variable in question. *Hypertension*: as derived from multiple administrative registers. *CAD*: coronary artery disease. *T2D*: Type 2 diabetes. *DF5*: FinnGen Data Freeze 5^7^. Stratification was based on self-reported sex.

| STUDY | VARIABLE | SEX-COMBINED | MEN | WOMEN |
| --- | --- | --- | --- | --- |
| SIGN | Sample size | 12,145 | 6,801 | 5,344 |
| | Age at onset (mean ± sd) in years | 67.6 ± 14.6 | 65.5 ± 13.8 | 70.2 ± 15.3 |
|  | Hypertension | 7,888 (65.0 %) | 4,269 (62.8 %) | 3,619 (67.8 %) |
|  | CAD | 2401 (19.8 %) | 1,438 (21.2 %) | 963 (18.0 %) |
|  | T2D | 2,757 (22.7 %) | 1,598 (23.5 %) | 1,159 (21.7 %) |
|  | Current smoker | 2,665 (22.0 %) | 1,715 (25.3 %) | 950 (17.8 %) |
| FINNGEN DF5 | Sample size | 8,124 | 4,708 | 3,416 |
| | Age at onset (mean ± sd) in years | 66.7 ± 13.3 | 66.0 ± 12.5 | 67.8 ± 14.3 |
|  | Hypertension | 5,274 (64.9%) | 3,017 (64.1%) | 2,257 (66.1%) |
|  | CAD | 2,582 (31.8%) | 1,775 (37.7%) | 807 (23.6%) |
|  | T2D | 2,648 (32.6%) | 1,645 (34.9%) | 1,003 (29.4%) |
|  | Current smoker | - | - | - |
| WHI | Sample size | - | - | 3,415 |
| | Age at onset (mean ± sd) in years | - | - | 76.6 ± 7.1 |
|  | Hypertension | - | - | 2,720 (79.65%) |
|  | CHD | - | - | 821 (24.04%) |
|  | T2D | - | - | 846 (24.77%) |
|  | Current smoker | - | - | 258 (7.55%) |

In the SiGN Discovery cohorts, genome-wide association analyses revealed 61 individual loci associated with stroke AAO in the sex-combined analysis at a significance threshold of p < 1.0×10^−5^ (see **Methods**), 144 individual loci in the men only analysis, and 37 individual loci in the women only analysis (**Figure S1, Figure S2**). There was one genome-wide significant hit (rs6051656) in the men-only analyses in SiGN spanning a 14 kb region at chromosome 20:364,295–378,978 near *TRIB3*, **Figure S3, Table S1**), which did not replicate in the men-only meta-analysis of SiGN with FinnGen (**Table S1**). Further meta-analyses of all SNPs associated at p < 1.0×10^−5^ in SiGN with FinnGen did not yield genome-wide significant results for the sex-combined analysis, but did reveal a genome-wide significant association in the women-only analysis with rs429358 on chromosome 19 at the Apolipoprotein E (*APOE*) locus (p_meta,SiGN-FinnGen_-value= 2.4×10^−8^, beta = -1.63 years ± 0.29, **Table 2, Figure S4**). This SNP was further replicated in WHI (p_meta,SiGN-FinnGen-WHI_ = 2.48×10^−11^, beta = -1.29 ± 0.38). The APOE-rs429358 is associated with stroke AAO in both sexes combined, although the magnitude of association is stronger in women than men (unequal variances t-test, p = 4.3×10^−4^, **Figure 1**). Conditional analysis (using the SiGN summary statistics) indicated no secondary associated SNPs at this locus.

**Table 2.** Meta-analysis results, total sample and sex-specific, of the association of *APOE*-rs429358 (chr19:45,411,941, b37) with ischemic stroke age of onset in Discovery (SiGN) and Replication (FinnGen and WHI) cohorts. The *sample size* for total sample and for men/women. The *alleles* are the effect (C) allele and other (T) allele, respectively, with the corresponding effect allele frequency (*EAF*). The *beta* (effect size) and the *se* (standard error) corresponds to difference in age of onset (in years) associated with each copy of the risk allele (C). As WHI includes only women, results presented for ‘Women-only’.

| Group | Discovery |  |  |  | Replication |  |  |  |  |  |  |  | Meta-analysis |  |  |  |
| --- | --- | --- | --- | --- | --- | --- | --- | --- | --- | --- | --- | --- | --- | --- | --- | --- |
|  | SiGN |  |  |  | FinnGen |  |  |  | WHI |  |  |  | (n = 22,126; 10,886/11,510) |  |  |  |
|  | (n = 10,857; 6,178/4,679) |  |  |  | (n = 8,124; 4,708/3,416) |  |  |  | (n = 3,415; 0/3,415) |  |  |  |  |  |  |  |
|  | EAf | beta | se | p-value | EAf | beta | se | p-value | EAf | beta | se | p-value | EAf | beta | se | p-value |
| *Sex-combined* | 0.13 | -1.21 | 0.25 | 1.60x10<sup>-6</sup> | 0.18 | -0.53 | 0.26 | 0.04 | - | - | - | - | 0.16 | -0.88 | 0.18 | 1.19x10<sup>-6</sup> |
| *Men-only* | 0.14 | -0.80 | 0.32 | 0.01 | 0.18 | 0.13 | 0.32 | 0.68 | - | - | - | - | 0.16 | -0.34 | 0.23 | 0.14 |
| *Women-only* | 0.13 | -1.78 | 0.40 | 9.70x10<sup>-6</sup> | 0.17 | -1.47 | 0.43 | 5.70x10<sup>-4</sup> | 0.13 | -1.02 | 0.26 | 6.97x10<sup>-5</sup> | 0.14 | -1.29 | 0.19 | 2.48x10<sup>-11</sup> |

**Figure 1:**
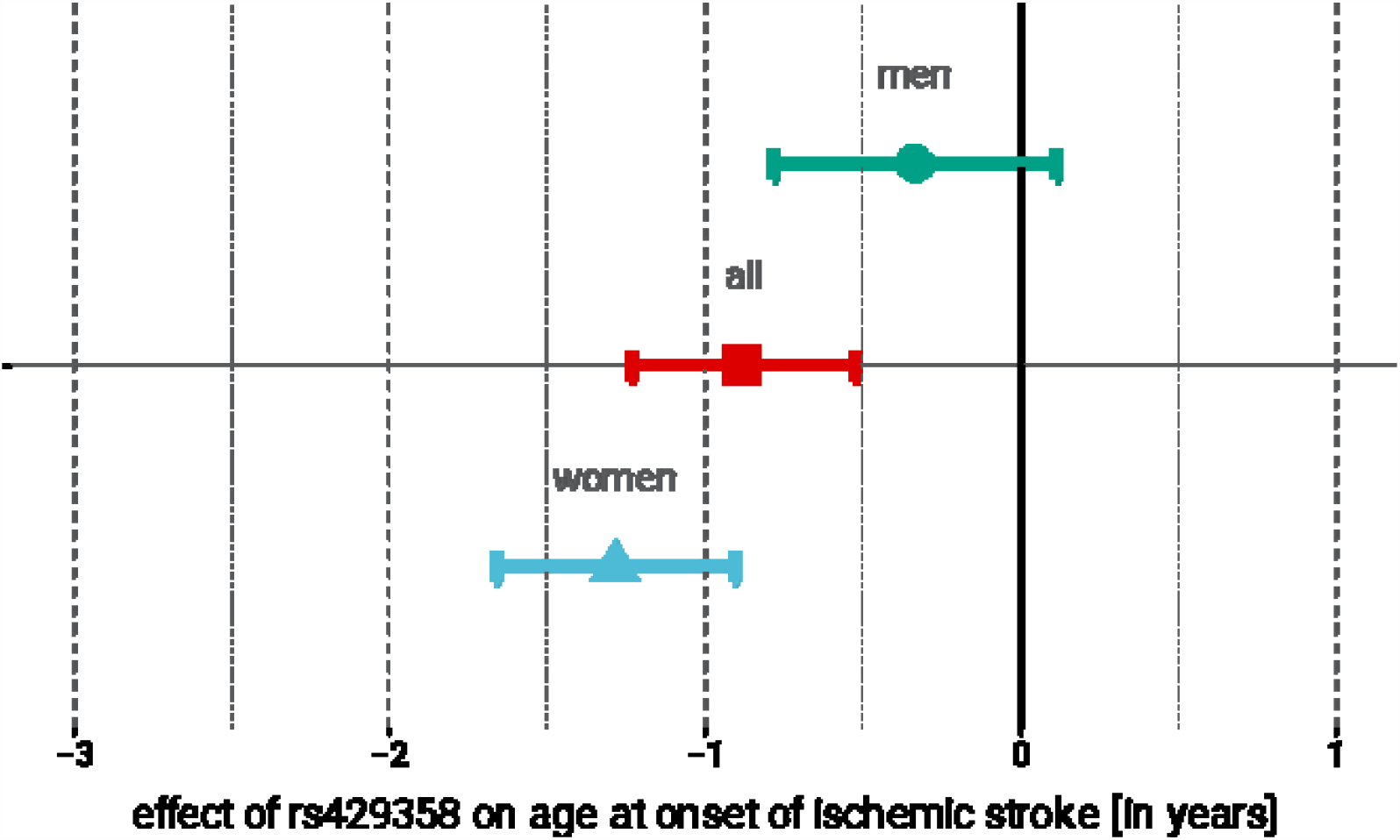
Association of APOE rs429358 with age of stroke onset in women (red), men (blue), and sex-combined meta-analysis. Point estimate represents effect of each copy of the minor (C) allele on AOO (in years); 95 % confidence intervals are indicated by error bars.

*APOE* rs429358 is a missense variant for which the minor allele, C, changes the amino acid at the 112th position of the ApoE protein from a cysteine to an arginine, thus altering the protein conformation. In combination with a second SNP (rs7412:C>T) in *APOE*, rs429358 encodes an individual’s ApoE isoform, with the rs429358-C allele associated with the ApoE-є4 allele. The ApoE-є4 allele (frequency of 0.16 in SiGN) has been associated with numerous adverse health outcomes, including hyperlipidemia^11^ and lipid metabolism^12^, Alzheimer’s disease^13^ and dementia, and coronary disease^14^. However, this variant is not associated with ischemic stroke susceptibility in MEGASTROKE (OR = 1.00; 95% CI: 0.96-1.03; p = 0.77), nor with any other ischemic stroke subtype (**Table S2**)^9^.

An association of *APOE* rs429358 with stroke AAO has been reported previously^3^ and this variant has also been associated with longevity^15^ and with age of parental death^10^. These observations, coupled with the association we observed between *APOE* rs429358 and stroke AAO, albeit genome-wide significant in women only, prompted us to investigate via simulation whether the stroke AAO association could be a manifestation of a survival bias attributable to a higher overall mortality among ApoE-є4 carriers.

We simulated a population of individuals who were followed from birth until death based on age-specific mortality rates obtained from the Social Security Administrations Actuarial Life Tables (**Figure 2**)^16^. Birthdates for the simulated subjects were randomly drawn between 1 January 1900 and 1 January 2020. Each individual was assigned a genotype for three SNPs, G_ISmultiplicative_, G_ISadditive_ and G_death_. G_ISmultiplicative_ and G_ISadditive_ increased risk of ischemic stroke only, and G_death_ increases risk of death only, but not through IS. Stroke was assumed to increase the risk of death as a function of the time since the event. We performed association analyses for each simulated SNP and two phenotypes: logistic regression of case-control status, and linear regression of age at onset. See **Methods** for simulation details.

**Figure 2:**
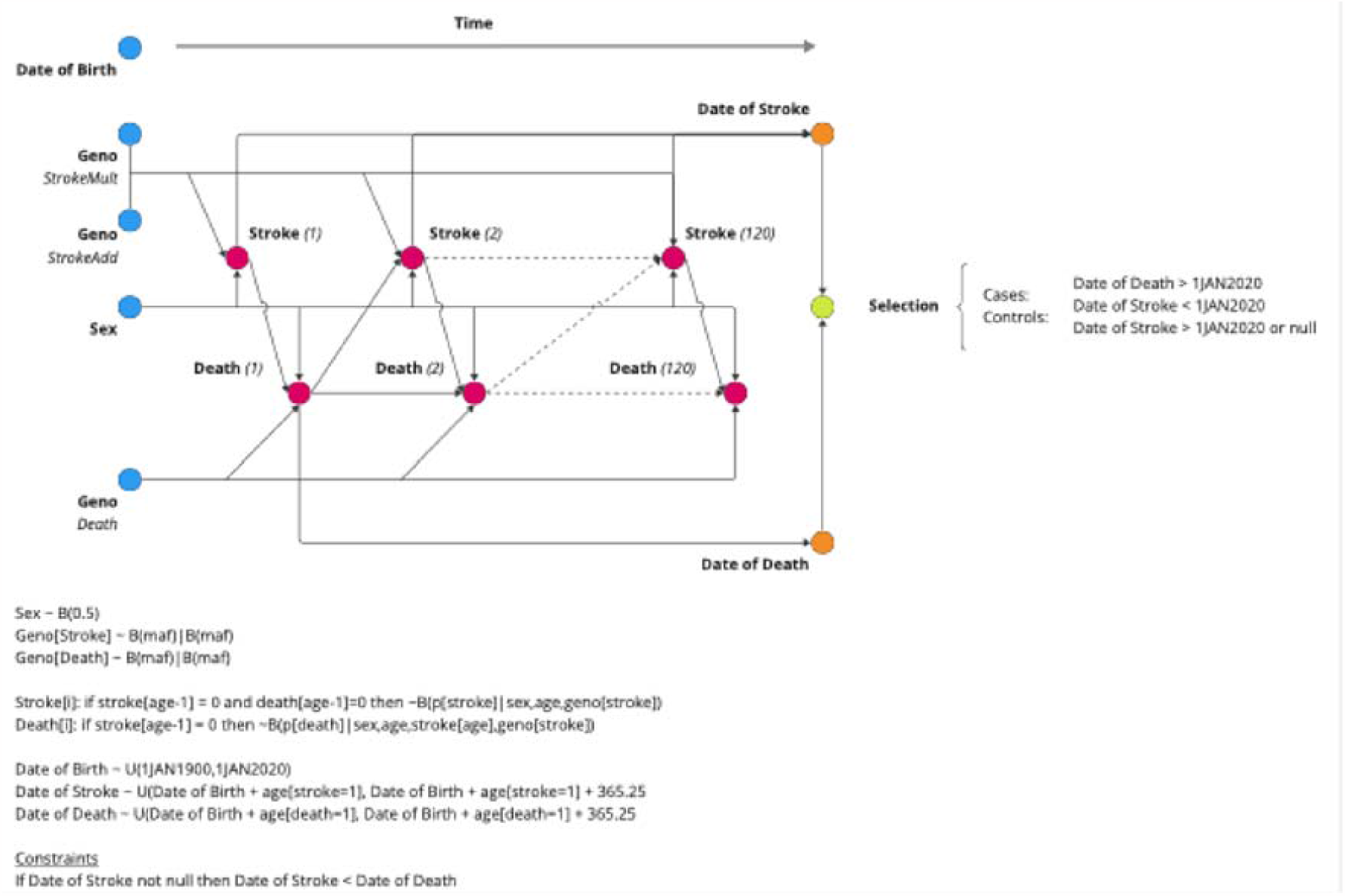
Data generating model for simulation study. Five variables were generated at birth (DOB, GENO and SEX) and subsequent risk of stroke and death were estimated annually. Date of death and date of stroke (if applicable) are outputted from the model.

As expected, both the additive and multiplicative loci simulated to influence risk of stroke were identifiable via a case-control design (1,000 cases and 1,000 controls), with power increasing with larger effect sizes (**Figure 3A**). Whether they were associated with AAO depended on the amount of risk conferred by the allele (**Figure 3B**). Loci with a relative increase in risk were not associated with AAO at all, but those with an additive increase in risk saw proportionally more stroke at early ages and thus the risk allele was associated with a lower AAO of IS. The simulated locus that was associated with mortality via mechanisms unrelated to stroke was not associated with stroke risk. However, that locus was associated with AAO. A locus with a two-fold increase in mortality would display an association with a ∼1.5 year decrease in age at onset, an effect size similar to that identified for the *APOE* locus in our GWAS. In other words, our results indicate that the observed SNP association in women is biased by an association with earlier death, assuming that the SNP’s effect is indeed independent of ischemic stroke.

**Figure 3:**
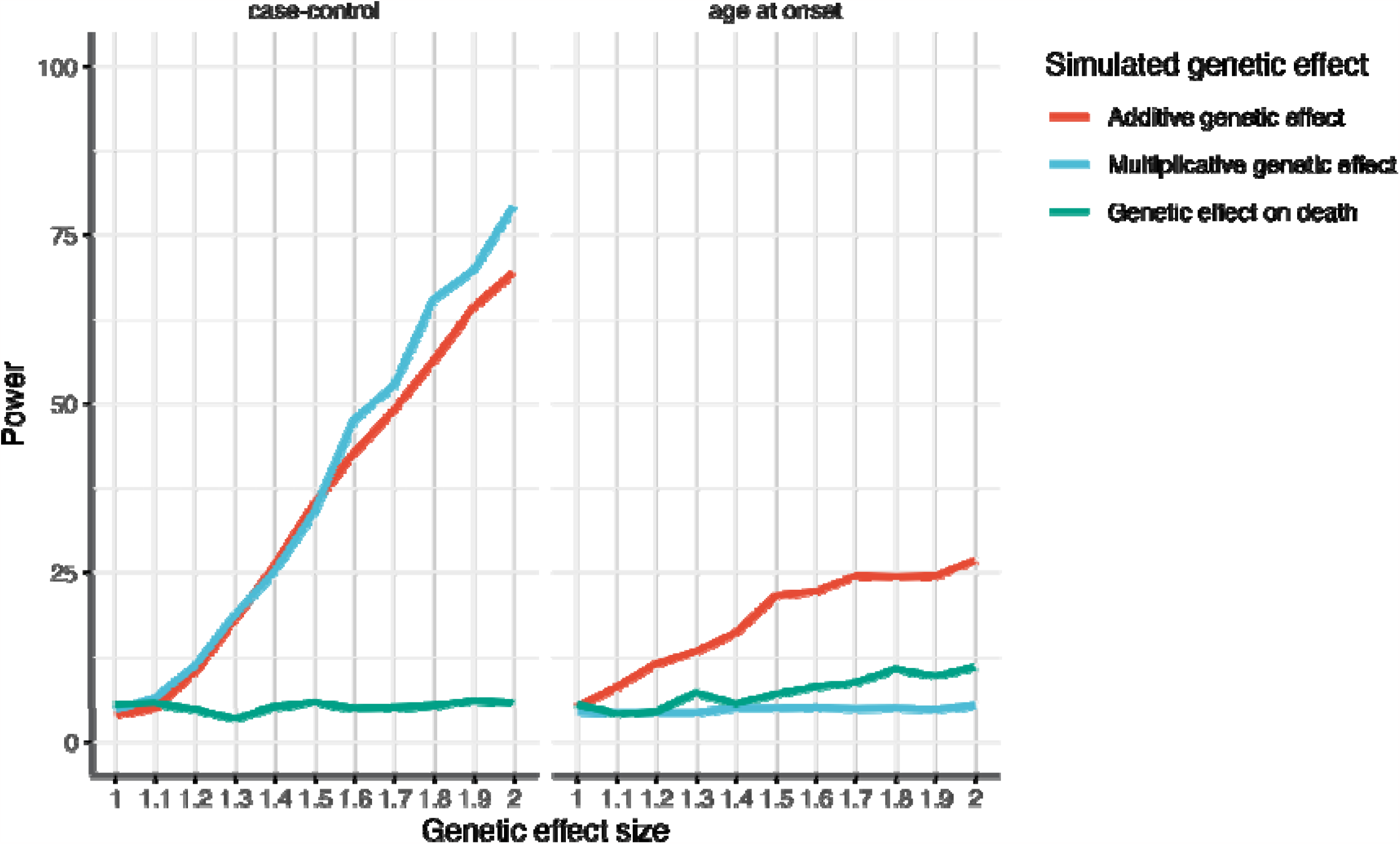
Estimated power to detect an association between genotype and stroke according to effect size for variants associated with age at death (green), additive effect on stroke susceptibility (red), and multiplicative effect on stroke susceptibility (blue) for (left) case-control analysis and (right) age at onset analysis. The x-axis shows the effect size in relative risk for a given genetic variant. The y-axis shows the power given a genetic effect size.

## Discussion

We found that *APOE*-rs429358, encoding the ApoE-є4 haplotype, is associated with earlier age of ischemic stroke in women. Although this SNP has not been associated with increased risk of ischemic stroke in prior GWAS, there are several lines of evidence supporting that the association we observed with stroke AAO is real. First, this association is statistically robust and replicated in two independent populations. Second, this SNP has been associated previously in a candidate gene study with stroke AAO^3^. Third, this SNP is biologically compelling as it encodes the ApoE-є4 allele, which has been associated previously with shorter lifespan^17–19^ as well as with several common age-related conditions, including Alzheimer’s disease^20^ and ischemic heart disease^20–23^.

While the association between rs429358 and ischemic stroke AAO appears to be robust, the interpretation of this association can be debated. One possibility is that this variant, while not associated with stroke susceptibility per se, is associated with the age at which stroke occurs in those more likely (either through genetic or environmental factors) to experience a stroke. A second possibility is that the association of this variant with stroke AAO is a more general consequence of the APOE risk allele at this locus being associated with shortened lifespan in general, *i*.*e*., an age-related decrease in the frequency of the risk allele. Our simulation study supports the latter interpretation. In other words, the association we detected between APOE and stroke AAO may be due to survival bias. Our simulations suggest that a locus exerting a 2-fold increase in mortality would also be associated with a ∼1.5 year decrease in age at onset, an effect size similar to that identified for the *APOE* locus in our GWAS and is consistent with previous findings^24,25^.

While previous simulation studies have been conducted assessing the power of AAO studies^26^, these previous studies have failed to account for the competing risk of mortality and potential bias that may play a role in these studies. Our simulations utilize a prospective data generating model, even though a retrospective case-control or case-only analytic model is fit to the data. This allows our simulations to estimate associations (either OR from a case-control study or years difference in AAO studies) that are more likely to represent those found in the natural population.

The reason for the predominantly larger effect on stroke AAO we observed among females compared to male stroke cases is not apparent. Possible explanations include a disproportionate effect of ApoE-є4 on survival in women compared to men, a direct effect of ApoE-є4 on stroke AAO, or chance. Modest differences in the associations of *APOE* genotype have been reported between men and women for ischemic heart disease^23^, Alzheimer’s Disease^27^, and lifespan^28^, although the reasons underlying these differences are not known. An alternative hypothesis may be more plausible and seems supported by our simulations and GWAS results. It is thought that ApoE-є4 is the proinflammatory ancestral allele in the human species and adaptive to reducing mortality under conditions of infections, food scarcity and (paradoxically) a shortened life expectancy^29^. Several lines of evidence support this notion. ApoE-є4 show less severe liver damage during hepatitis C infections, the allele frequency remained high in populations where food was scarce up on to recently, and the allele is associated with higher circulating cholesterol^30,31^. As the human population expanded and thrived, the ApoE-є3 and ApoE-є2 alleles spread, while the ApoE-є4 was maintained by balancing selection^25,32^. Yet, as conditions for our species continued to improve, diets and lifespan changed, thus rendering ApoE-є4 susceptible to CVD and more pronounced effects in women^25^.

Strengths of our study include the large number of well characterized ischemic stroke cases with a wide range of ages of onset as well as the prospective design of our simulation study. Nevertheless, our study is not without limitations. One notable result is that effect size in WHI women (−1.02) is the same direction but smaller in magnitude, possibly reflecting the fact these women, on average, experienced stroke onset at substantially older age with higher prevalence of hypertension and lower current smoking than the other female participants and the men. True as this may be, if anything the overall meta-analysis effect estimate is smaller, which does not preclude the main interpretation that results are consistent despite variations across studies. However, foremost among the limitations, none of the studies included in our meta-analysis were birth cohorts, so only cases who survived until the age of their recruitment are part of our GWAS. If case-fatality rates differed between early and late onset stroke, then variants associated specifically with early (or late) onset stroke could go undetected. Likewise, our study may be affected by an ascertainment bias, as cases were drawn from case-control studies, *i*.*e*. people were ascertained on phenotype which may introduce confounding such as some strokes that are more likely to be caught by clinicians or the severity of stroke. However, the replication in FinnGen sidesteps this issue because people are randomly ascertained in this study.

In conclusion, we have detected a robust association of the *APOE* locus with stroke AAO and provide simulations to suggest that this association may be unrelated to ischemic stroke per se but related to a general survival bias.

## Data Availability

Summary statistics from the combined, women-only, and men-only discovery GWAS in SiGN are available through dataverseNL and GWAS Catalog.

## Acknowledgements

We would like to thank Sara L. Pulit for supervision and mentoring during initiation of this study.

We want to acknowledge the participants and investigators of FinnGen study. Following biobanks are acknowledged for delivering biobank samples to FinnGen: Auria Biobank (www.auria.fi/biopankki), THL Biobank (www.thl.fi/biobank), Helsinki Biobank (www.helsinginbiopankki.fi), Biobank Borealis of Northern Finland (https://www.ppshp.fi/Tutkimus-ja-opetus/Biopankki/Pages/Biobank-Borealis-briefly-in-English.aspx), Finnish Clinical Biobank Tampere (www.tays.fi/en-US/Research_and_development/Finnish_Clinical_Biobank_Tampere), Biobank of Eastern Finland (www.ita-suomenbiopankki.fi/en), Central Finland Biobank (www.ksshp.fi/fi-FI/Potilaalle/Biopankki), Finnish Red Cross Blood Service Biobank (www.veripalvelu.fi/verenluovutus/biopankkitoiminta), Terveystalo Biobank (www.terveystalo.com/fi/Yritystietoa/Terveystalo-Biopankki/Biopankki/) and Arctic Biobank (https://www.oulu.fi/en/university/faculties-and-units/faculty-medicine/northern-finland-birth-cohorts-and-arctic-biobank). All Finnish Biobanks are members of BBMRI.fi infrastructure (www.bbmri.fi). Finnish Biobank Cooperative FINBB (https://finbb.fi/) is the coordinator of BBMRI-ERIC operations in Finland.

## Data and code availability

Genotypes and phenotypes for the SiGN cases are available on dbGAP under accession number: phs000615.v1.p1. The Finnish biobank data can be accessed through the Fingenious® services (https://site.fingenious.fi/en/) managed by FINBB. WHI data is available through dbGaP through accession number phs000120.v12.p3. Summary statistics from the combined, women-only, and men-only discovery GWAS in SiGN are available through GWAS Catalog. The scripts used for these analyses and for the preparation for GWAS Catalog are available on GitHub: https://github.com/CirculatoryHealth/AAO_IschemicStroke.

## Methods

### Genome-wide analysis of stroke age at onset

#### Study populations

Analyses were performed on subjects of European ancestry from the Stroke Genetics Network (SiGN), the FinnGen Study and the Women’s Health Initiative (WHI). SiGN is an international collaboration that includes ischemic stroke cases recruited from multiple sites in the United States and Europe (UK, Poland, Belgium, Spain, Austria, and Sweden)^6^. The Discovery phase of this analysis included 10,857 SiGN participants, the Replication phase included 8,124 participants from FinnGen Data Freeze 5^7^, and 3,415 participants from the Women’s Health Initiative (WHI)^8^. Participants provided (written) informed consent and all studies upheld the ethical standards according to the Helsinki Declaration.

#### Phenotype definition

For each study included in the SiGN dataset, ischemic stroke was confirmed by neuroimaging. Details, including inclusion criteria, for each study can be found in the original SiGN GWAS publication^32^. In WHI all incident strokes, other vascular events, and deaths were identified through self-report at annual (OS) and semi-annual (CT) participant contacts, and through third party reports by family members and proxies. Medical records were obtained for potential strokes, and adjudication was performed by trained physician adjudicators who assigned a diagnosis. Stroke diagnosis requiring and/or occurring during hospitalization was based on rapid onset of a neurological deficit attributable to an obstruction or rupture of an arterial vessel system. The deficit was not known to be secondary to brain trauma, tumor, infection or other cause and must have lasted more than 24 hours unless death supervened or a lesion compatible with acute stroke was evident on computed tomography or magnetic resonance imaging scan^33^. Strokes were classified as ischemic, hemorrhagic, or unknown/missing. Ischemic stroke subtypes were further classified using Trial of Org 10172 in Acute Stroke Treatment (TOAST) criteria^34^.

#### DNA isolation, genotyping, and imputation

The SiGN and WHI participants have been genotyped on different Illumina platforms. The dataset was split in different study strata, based on similar genetic ancestry and genotyping platforms. Genotypes that were not measured were imputed against 1000G phase 3 for SiGN^32^, and TOPMed for WHI^35^. The FinnGen cohort and methods are comprehensively described elsewhere^7^.

#### Genome-wide linear regression

Genome-wide analyses of stroke AAO were carried out in SiGN using BOLT-LMM^37^. We used PLINK 1.9^36^ to ‘hard call’ and subset imputed SNPs to build the genetic relationship matrix (GRM). SNPs with an INFO < 0.8, genotyping rate < 95%, or missing genotype rate > 5% were excluded from analysis. Covariates in the association analysis included study stratum, the GRM to correct for the subtle population stratification, and sex (in the sex-combined analysis). Post analysis, we filtered out SNPs with minimum minor allele frequency (MAF) < 5%.

#### Independent signals

To identify independent signals for the three main analyses in the Discovery we used PLINK 1.9 with 1000G phase 1 (version 3) as a reference^37^. We set the minimal p-value threshold at 0.05, defined a clumped region as ±500kb, with a minimum linkage disequilibrium r^2^ at 0.05 `(--clump-p1 5e-8 --clump-p2 0.05 --clump-kb 500 --clump-r2 0.05 --clump-best --clump-verbose`). To identify independent signals to take forward in the Replication phase, we include clumps at p < 1.0×10^−5^ `(-- clump-p1 1e-5 --clump-p2 0.05 --clump-kb 500 --clump-r2 0.05 -- clump-best --clump-verbose`).

#### Conditional analysis

We used GCTA COJO^38^ to investigate whether there were additional associated SNPs at the discovered loci. We used the stepwise model selection procedure (`--cojo-slct`) and used the imputed genotype data (converted to ‘hard call’, as described for the GWAS) as input (`--bfile`).

#### Meta-analysis

We performed a look-up of SNPs that were associated at p < 1.0×10^−5^ in any of the three analyses (sex-combined, men-only, women-only) in FinnGen and in the Women’s Health Inititiave (women-only, and for rs429358 only)^8^. Meta-analyses of SiGN with FinnGen were performed in METAL using the inverse variance weighted approach^39^. We considered a p-value threshold of p < 5 × 10^−8^ to be significant in the meta-analysis. Baseline characteristics for the replication datasets can be found in Table 1.

#### Testing for sex differences in effect size

To test how likely the differences in effect size between the women and men analysis are, under the null hypothesis of no difference, we used a t-test for unequal variance. The test we used is similar to Welch’s t-test, but we additionally correct for the Spearman rank correlation *r* between all women and men effect sizes (filtered on MAF > 0.05 and INFO > 0.8)^40^; *r* was equal to 0.018.

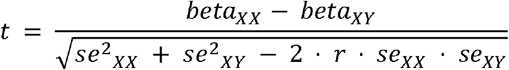

### Simulations

#### Data generating model

The data generating model is presented in Figure 1. Pseudo-men and women were simulated drawing a date of birth at random from 1 January 1900 to 1 January 2020. Each pseudo-individual was followed over the course of 120 years or until their death, whichever came first. At birth, genotypes were assigned at three loci each having a minor allele frequency of 10%. Two genotypes, Geno_StrokeMult_ and Geno_StrokeAdd_ incurred a risk on stroke only, and the other, Geno_Death_ incurred a risk on death via an unspecified pathway independent of stroke. The annual stroke risk was a function of sex, age and genotypes given by:

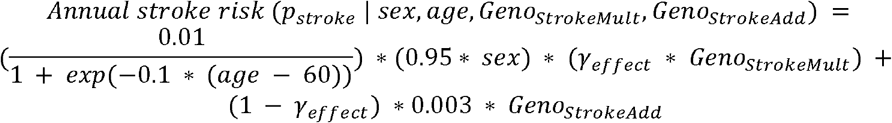

The genetic effect of the genotype, *γ*_*effect*_, was simulated from 1.0 to 2.0 in increments of 0.1. An initial stroke event was drawn from a Bernoulli distribution with probability (*p*_*stroke*_ | *sex, age, Geno*_*StrokeMult*_, *Geno*_*StrokeAdd*_) given that the subject had not died previously and had not previously experienced a stroke. If the binomial draw indicated a stroke at that age, an exact date of stroke was randomly drawn from a uniform distribution of days in that year. Baseline annual risk of death was taken from the Social Security Administrations Actuarial Life Tables^16^. The mortality effect of Geno_Death_ was simulated using the same range of parameters, *γ*_*effect*_, and was a function of age. The relative increase in risk was assumed to be close to null at young ages and then increased over the lifetime until a pre-specified risk ratio. Stroke was assumed to increase the risk of death as a function of the time since the event, given by

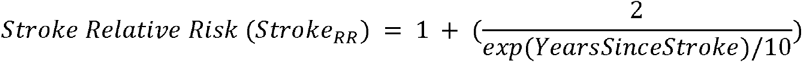

The resulting annual mortality risk was given by

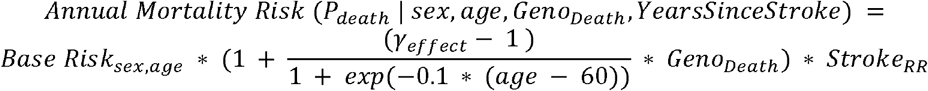

The given data generating model resulted in observations with 7 features: date of birth, sex, *Geno*_*StrokeMult*_, *Geno*_*StrokeAdd*_, Geno_Death_, date of stroke, and date of death. Random draws of pseudo-individuals were made from the data generating model who were (1) alive as of 1 Jan 2020 and (2) over the age of 18 on that date until 1,000 cases (defined as having a stroke prior to 1 Jan 2020) and 1,000 controls (defined as never having a stroke or having a stroke after 1 Jan 2020) were drawn. Each simulation scenario was replicated 1,000 times to make robust estimates of the mean of estimated parameters and standard errors. The simulation study was performed by using SAS (v9.4).

#### Genotypic models

Two genotypic models were simulated. The first modeled a constant relative risk over the lifespan, given by *γ*_*effect*_ and parameterized as a risk ratio. The second modeled a constant additive risk over the lifespan given by a function of *γ*_*effect*_ as shown above. This model simulated a larger relative effect at younger ages than at older ages. It has been hypothesized that some genetic loci may have a disproportionate effect on stroke risk at younger ages versus older ages, and thus genetic contributors to stroke risk may be easier to find^7^. For example, when *γ*_*effect*_ = 1.1, the early onset locus had a relative risk of 1.6 at age 30, 1.1 at age 50 and 1.04 at age 70. This allows for a test of the ability of age at onset analyses to identify loci that have a larger relative effect early in life rather than later.

#### Target Parameter

Given the above data generating model, it is trivial to determine the age at stroke for each pseudo-individual (date of stroke – date of birth). The target parameter was defined as the difference in the age of stroke between genotypes among cases.

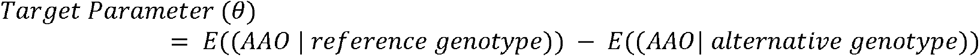

Estimates of this target parameter were made using linear regression controlling for sex to approximate a common GWAS strategy. Genotypes were coded as 0,1,2 to estimate the additive genetic model. Models were run for each of the simulated loci separately.

## Ethics statement

Patients and control subjects in FinnGen provided informed consent for biobank research, based on the Finnish Biobank Act. Alternatively, separate research cohorts, collected prior the Finnish Biobank Act came into effect (in September 2013) and start of FinnGen (August 2017), were collected based on study-specific consents and later transferred to the Finnish biobanks after approval by Fimea, the National Supervisory Authority for Welfare and Health. Recruitment protocols followed the biobank protocols approved by Fimea. The Coordinating Ethics Committee of the Hospital District of Helsinki and Uusimaa (HUS) approved the FinnGen study protocol Nr HUS/990/2017.

The FinnGen study is approved by Finnish Institute for Health and Welfare (THL), approval number THL/2031/6.02.00/2017, amendments THL/1101/5.05.00/2017, THL/341/6.02.00/2018, THL/2222/6.02.00/2018, THL/283/6.02.00/2019,

THL/1721/5.05.00/2019, Digital and population data service agency VRK43431/2017-3, VRK/6909/2018-3, VRK/4415/2019-3 the Social Insurance Institution (KELA) KELA 58/522/2017, KELA 131/522/2018, KELA 70/522/2019, KELA 98/522/2019, and Statistics Finland TK-53-1041-17.

The Biobank Access Decisions for FinnGen samples and data utilized in FinnGen Data Freeze 5 include: THL Biobank BB2017_55, BB2017_111, BB2018_19, BB_2018_34, BB_2018_67, BB2018_71, BB2019_7, BB2019_8, BB2019_26, Finnish Red Cross Blood Service Biobank 7.12.2017, Helsinki Biobank HUS/359/2017, Auria Biobank AB17-5154, Biobank Borealis of Northern Finland_2017_1013, Biobank of Eastern Finland 1186/2018, Finnish Clinical Biobank Tampere MH0004, Central Finland Biobank 1-2017, and Terveystalo Biobank STB 2018001.

