## Supplemental Material for "Evidence of survival bias in the association between *APOE-Є4* and age of ischemic stroke onset"

accompanying

### Supplemental Figures

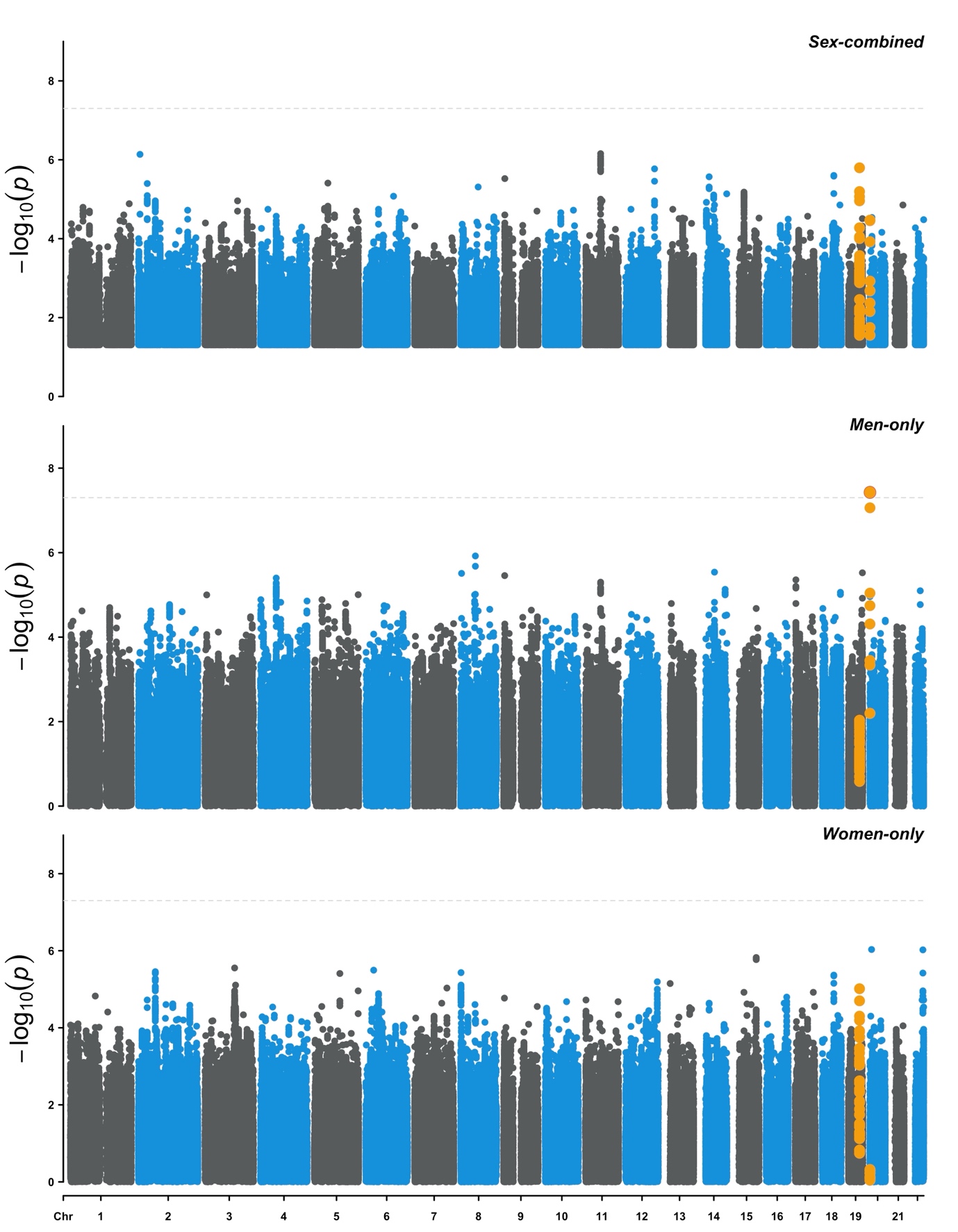

**Figure S1. Manhattan plots in the Discovery phase.** X-axis shows the chromosome base pair position. Y-axis shows the -log_10_ of the p-value of association with age at ischemic stroke onset. *Upper panel:* Manhattan plot of age at onset in sex-combined European IS cases. *Middle panel:* Manhattan plot of age at onset in men-only European IS cases. *Lower panel:* Manhattan plot of age at onset in women-only European IS cases. Highlighted (golden) are variants in LD with the rs429358 (r^2^ > 0.1) on chromosome 19 near *APOE*, and rs6051656 on chromosome 20 near *TRIB3*.

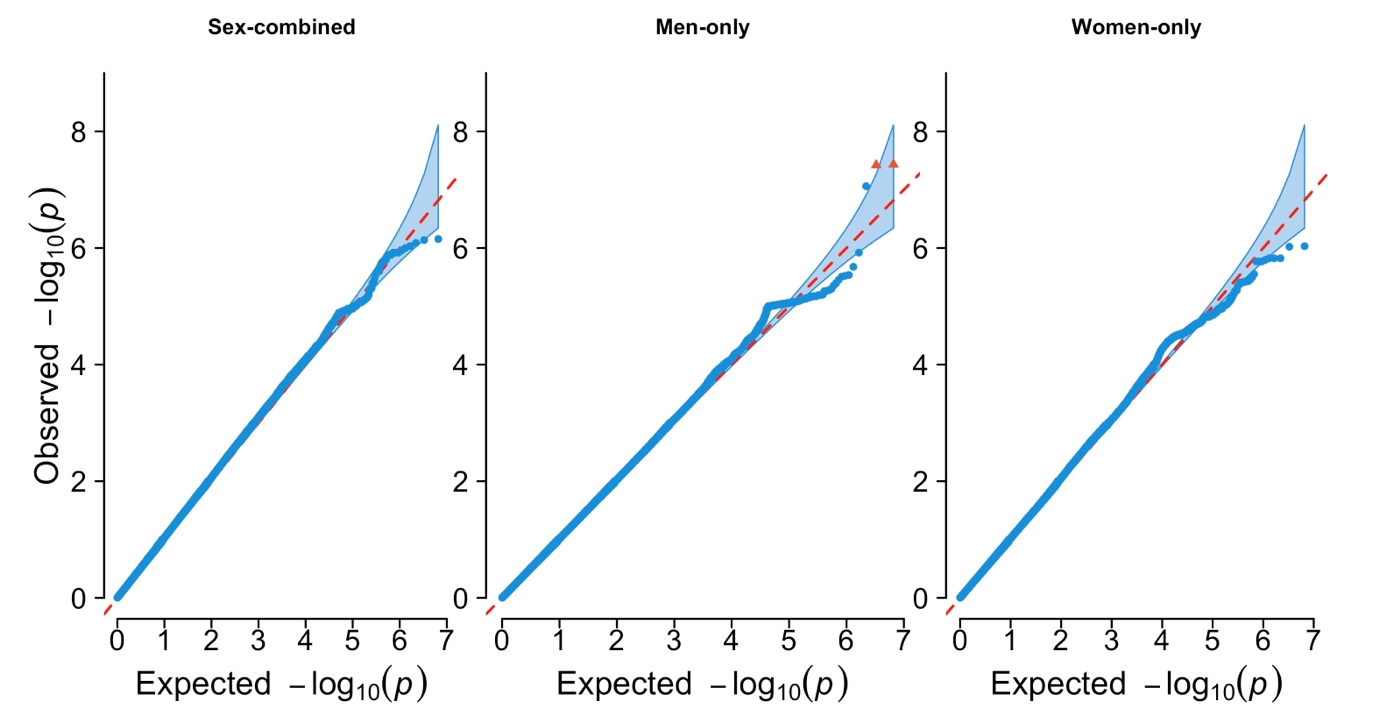

**Figure S2. QQ plots in the Discovery phase.** X-axis shows the expected -log_10_ of the p-value of association with age at ischemic stroke onset, whereas the y-axis shows the observed -log_10_ p-values. *Upper panel:* QQ plot of age at onset in sex-combined European IS cases; genomic inflation factor λ = 1.05. *Middle panel:* QQ plot of age at onset in men-only European IS cases; genomic inflation factor λ = 1.00. *Lower panel:* QQ plot of age at onset in women-only European IS cases; genomic inflation factor λ = 1.05.

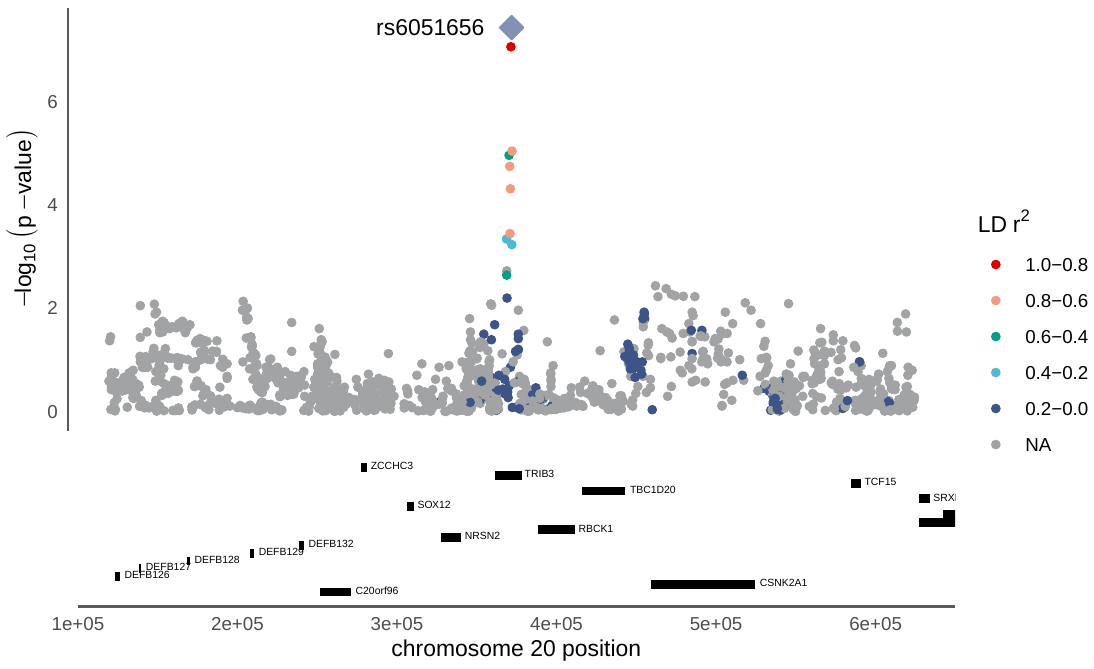

**Figure S3. Regional association plot in the *TRIB3* locus in the Discovery phase.** X-axis shows the chromosome base pair position, and the genes (black). The y-axis shows the -log_10_ of the p-value of association with age at onset of ischemic stroke. Colors indicate the linkage disequilibrium r^2^ relative to the lead variant (purple).

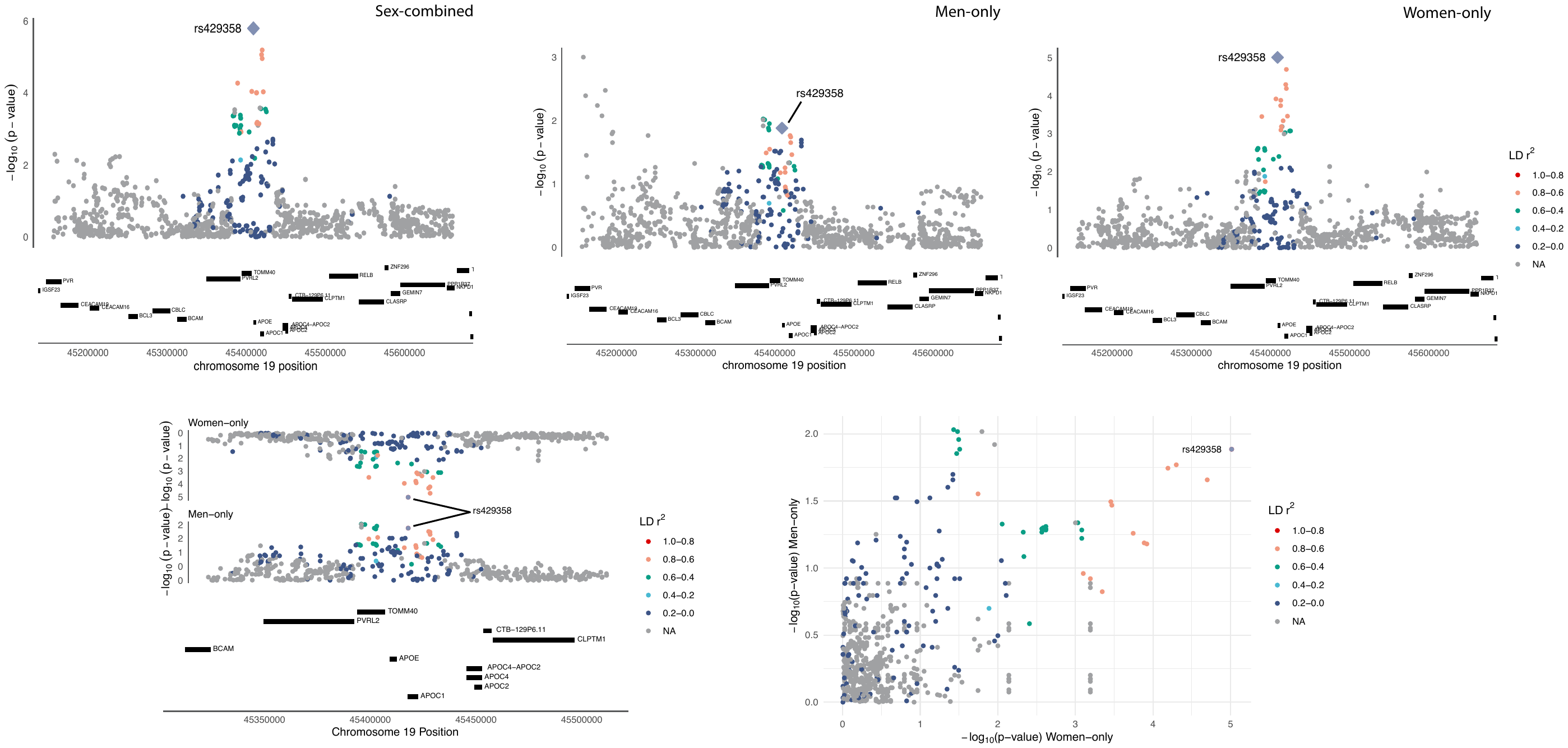

**Figure S4. Regional association plot in the *APOE* locus across analyses in the Discovery phase.** *Top-panel.* X-axis shows the chromosome base pair position, and the genes (black). The y-axis shows the -log_10_ of the p-value of association with age at onset of ischemic stroke. *Lower panel, left.* Mirror plot comparing the results from the men- and women-only analyses in a 100,000 bp region around rs429358. *Lower panel, right.* Scatter plot comparing the associated -log_10_ of the p-values of association with age at onset of ischemic stroke between the sexes. Colors indicate the linkage disequilibrium r^2^ relative to the lead variant (purple).

### Supplemental Tables

**Table S1. Meta-analysis results of the association of rs6051656 and rs429358 with ischemic stroke age of onset in Discovery (SiGN) and FinnGen cohorts.** The sample size for total sample and for men/women. The *alleles* are the effect and other allele, respectively, with the corresponding effect allele frequency (EAF). The *beta* (effect size) and the *se* (standard error) corresponds to difference in age of onset (in years) associated (*p-value*) with each copy of the effect allele.

| **Group** | **SNP information** | | | | **Discovery** | | | | **Replication** | | | | **Meta-analysis** | | | |
| --- | --- | --- | --- | --- | --- | --- | --- | --- | --- | --- | --- | --- | --- | --- | --- | --- |
|  |  | | | | **SiGN** | | | | **FinnGen** | | | | (n = 18,981; 10,886/8,095) | | | |
|  |  | | | | (n = 10,857; 6,178/4,679) | | | | (n = 8,124; 4,708/3,416) | | | |  | | | |
|  | **Locus** | **rsID** | **Chr:BP** | **Alleles** | **EAF** | **beta** | **se** | **p-value** | **EAF** | **beta** | **se** | **p-value** | **EAF** | **beta** | **se** | **p-value** |
| *Sex-combined* | *Trib3* | rs6051656 | chr20:374763 | C/T | 0.87 | 1.15 | 0.28 | 3.5x10-5 | 0.89 | 0.25 | 0.32 | 0.43 | 0.88 | 0.76 | 0.21 | 2.73x10^-4^ |
| *Men-only* |  |  |  |  | 0.86 | 1.95 | 0.35 | 3.7x10-8 | 0.89 | 0.50 | 0.52 | 0.33 | 0.88 | 1.14 | 0.26 | 1.75x10^-5^ |
| *Women-only* |  |  |  |  | 0.87 | 0.24 | 0.44 | 0.59 | 0.89 | 0.11 | 0.40 | 0.79 | 0.88 | 0.35 | 0.34 | 0.2997 |
| *Sex-combined* | *ApoE* | rs429358 | chr19:45411941 | C/T | 0.13 | -1.21 | 0.25 | 1.60x10^-6^ | 0.18 | -0.53 | 0.26 | 0.04 | 0.16 | -0.88 | 0.18 | 1.19x10^-6^ |
| *Men-only* |  |  |  |  | 0.14 | -0.80 | 0.32 | 0.01 | 0.18 | 0.13 | 0.32 | 0.68 | 0.16 | -0.34 | 0.23 | 0.14 |
| *Women-only* |  |  |  |  | 0.13 | -1.78 | 0.40 | 9.70x10^-6^ | 0.17 | -1.47 | 0.43 | 5.70x10^-4^ | 0.15 | -1.63 | 0.29 | 2.37x10^-8^ |

**Table S2. Meta-analysis results of the association of APOE-rs429358(chr19:** **45,411,941, b37) with ischemic stroke subtypes in MEGASTROKE.** The sample sizes are for the sex-combined analyses, sex-stratified is not available. The alleles are the effect (C) allele and other (T) allele, respectively, with the corresponding effect allele frequency (EAF). The *beta* (effect size) and the *se* (standard error) corresponds to the difference in risk associated (*p-value*) with each copy of the risk allele (C). The direction is the direction of the variant for the trait in each cohort contributing to MEGASTROKE (please refer to the original paper for a list of cohorts). The heterogeneity statistic is given (*Het I^2^*) with the Chi-squared (*Het Chi^2^*), and the degrees of freedom (Het Df) as well as the p-value (Het Pval). The *TotalSampleSize* indicates the sum of samples sizes of each contributing cohort in MEGASTROKE for each trait, along with the total sample size of the cases (*TotalCases*). These data are from an analysis of MEGASTROKE without SiGN cohorts and in populations of European-ancestry only.

| **Subtype** | **EAF** | **beta** | **se** | **p-value** | **direction** | **Het I^2^** | **Het Chi^2^** | **Het Df** | **Het Pval** | **TotalSampleSize** | **TotalCases** |
| --- | --- | --- | --- | --- | --- | --- | --- | --- | --- | --- | --- |
| AS | 0.15 | 0.020 | 0.01 | 0.18 | --+--+----+++-+- | 39.3 | 24.7 | 15 | 0.05 | 414,547 | 32,475 |
| IS | 0.15 | 0.005 | 0.02 | 0.77 | -+--+----+++-+- | 34.3 | 21.3 | 14 | 0.09 | 399,337 | 26,193 |
| CES | 0.15 | -0.031 | 0.03 | 0.35 | +++-+--+?+-+ | 0.0 | 8.4 | 10 | 0.59 | 289,083 | 4,908 |
| LAS | 0.15 | -0.003 | 0.04 | 0.94 | +--+++--- | 41.4 | 13.6 | 8 | 0.09 | 185,768 | 3,120 |
| SVS | 0.15 | -0.017 | 0.04 | 0.68 | -?+-+--+++- | 6.1 | 9.6 | 9 | 0.39 | 225,764 | 3,821 |

### Supplemental Text

#### Stroke Genetics Network Funding and Acknowledgments

The Stroke Genetics Network (SiGN) study was funded by a cooperative agreement grant from the National Institute of Neurological Disorders and Stroke (NINDS) U01 NS069208. Genotyping services were provided by the Johns Hopkins University Center for Inherited Disease Research (CIDR), which is fully funded through a federal contract from the National Institutes of Health (NIH) to the Johns Hopkins University (contract No. HHSN268200782096C). The Biostatistics Department Genetics Coordinating Center at the University of Washington (Seattle) provided more extensive quality control of the genotype data through a subcontract with CIDR. Additional support to the Administrative Core of SiGN was provided by the Dean’s Office, University of Maryland School of Medicine. Additional support for this study was also provided by P30 AG028747.

#### Funding per SiGN cohort

**ASGC:** Australian population control data were derived from the Hunter Community Study. We also thank the University of Newcastle for funding and the men and women of the Hunter region who participated in this study. This research was funded by grants from the Australian National and Medical Health Research Council (NHMRC Project Grant ID: 569257), the Australian National Heart Foundation (NHF Project Grant ID: G 04S 1623), the University of Newcastle, the Gladys M Brawn Fellowship scheme, and the Vincent Fairfax Family Foundation in Australia. Elizabeth G Holliday is supported by the Australian NHMRC Fellowship scheme.

**BASICMAR:** The Base de Datos de Ictus del Hospital del Mar (BASICMAR) Genetic Study was supported by the Ministerio de Sanidad y Consumo de España, Instituto de Salud Carlos III (ISC III) with the grants: Registro BASICMAR Funding for Research in Health (PI051737); GWA Study of Leukoaraiosis (GWALA) project from Fondos de Investigación Sanitaria ISC III (PI10/02064) and (PI12/01238); Agència de Gestió Ajuts Universitaris de Recerca (2014 SGR 1213) and Fondos European Regional Development Funding (FEDER/EDRF) Red de Investigación Cardiovascular (RD12/0042/0020). Additional support was provided by the Fundació la Marató TV3 with the grant GODS project. Genestroke Consortium (76/C/2011) Recercaixa’13 (JJ086116). Assistance with data cleaning was provided by the Research in Cardiovascular and Inflammatory Diseases Program of Institute Hospital del Mar of Medical Investigations, Hospital del Mar, and the Barcelona Biomedical Research Park.

**BRAINS:** The Bio-Repository of DNA in Stroke (BRAINS) was supported by the British Council (UKIERI), Henry Smith Charity, the UK Stroke Research Network and the Qatari National Research Fund. Prof Sharma was supported by a Department of Health (United Kingdom) Senior Fellowship.

**EDINBURGH:** The Edinburgh Stroke Study was supported by the Wellcome Trust and the Binks Trust. Sample processing occurred in the Genetics Core Laboratory of the Wellcome Trust Clinical Research Facility, Western General Hospital, Edinburgh, UK. Much of the neuroimaging occurred in the Scottish Funding Council Brain Imaging Research Centre (www.sbirc.ed.ac.uk), University of Edinburgh, a core area of the Wellcome Trust Clinical Research Facility and part of the Scottish Imaging Network–A Platform for Scientific Excellence (SINAPSE) collaboration (www.sinapse.ac.uk), funded by the Scottish Funding Council and the Chief Scientist Office. Genotyping was performed at the Wellcome Trust Sanger Institute in the United Kingdom and funded by the Wellcome Trust as part of the Wellcome Trust Case Control Consortium 2 project (085475/B/08/Z and 085475/Z/08/Z and WT084724MA).

**GCNKSS:** The Greater Cincinnati/Northern Kentucky Stroke Study (GCNKSS) was supported by the NIH (NS030678).

**GRAZ:** The Austrian Stroke Prevention Study was supported by the Austrian Science Fund (FWF) grant Nos. P20545-P05 and P13180 and I904-B13 (Era-Net). The Medical University of Graz supports the databases of the Graz Stroke Study and the Austrian Stroke Prevention Study.

**ISGS and SWISS:** The Ischemic Stroke Genetics Study (ISGS) was supported by the NINDS (R01 NS42733; PI Dr Meschia). The Sibling with Ischemic Stroke Study (SWISS) was supported by the NINDS (R01 NS39987; PI Dr Meschia). Both SWISS and ISGS received additional support, in part, from the Intramural Research Program of the National Institute on Aging (Z01 AG000954-06; PI Andrew Singleton). SWISS and ISGS used samples and clinical data from the NIH-NINDS Human Genetics Resource Center DNA and Cell Line Repository (http://ccr.coriell.org/ninds), human subject protocol Nos. 2003-081 and 2004-147. SWISS and ISGS used stroke-free participants from the Baltimore Longitudinal Study of Aging (BLSA) as controls with the permission of Dr Luigi Ferrucci. The inclusion of BLSA samples was supported, in part, by the Intramural Research Program of the National Institute on Aging (Z01 AG000015-50), human subject protocol No. 2003-078. This study used the high-performance computational capabilities of the Biowulf Linux cluster at the NIH (http://biowulf.nih.gov). For SWISS and ISGS cases of African ancestry, a subset of the Healthy Aging in Neighborhoods of Diversity across the Life Span study (HANDLS) were used as stroke-free controls. HANDLS is funded by the National Institute of Aging (1Z01AG000513; PI Michele K. Evans).

**KRAKOW:** Phenotypic data and genetic specimens collection were funded by the grant from the Polish Ministry of Science and Higher Education for Leading National Research Centers (KNOW) and by the grants from the Jagiellonian University Medical College in Krakow, Poland: K/ZDS/002848, K/ZDS/003844.

**LEUVEN:** The Leuven Stroke genetics study was supported by personal research funds from the Department of Neurology of the University Hospitals Leuven. Dr Thijs is supported by a Fundamental Clinical Research grant from FWO Flanders (Nos. 1800908N and 1800913N). Dr Lemmens is a Senior Clinical Investigator of FWO Flanders (FWO 1841913N) and is supported through Fonds Annie Planckaert-Dewaele. An Goris is supported by the Research Fund KU Leuven (OT/11/087) and Research Foundation Flanders (G073415N).

**LUND:** The Lund Stroke Register was supported by the Swedish Research Council (K2010-61X-20378-04-3), The Swedish Heart-Lung Foundation, Region Skåne, Skåne University Hospital, the Freemasons Lodge of Instruction EOS in Lund, King Gustaf V’s and Queen Victoria’s Foundation, Lund University, and the Swedish Stroke Association. Biobank services were provided by Region Skåne Competence Centre (RSKC Malmö), Skåne University Hospital, Malmö, Sweden, and Biobank, Labmedicin Skåne, University and Regional Laboratories Region Skåne, Sweden.

**MALMӦ:** The Malmӧ Diet and Cancer Study was supported by the Swedish Research Council (Vetenskapsrådet), Heart and Lung Foundation (Hjärt och Lungfonden), and Swedish Stroke Foundation (Strokeförbundet).

**MCISS:** The Middlesex County Ischemic Stroke Study (MCISS) was supported by intramural funding from the New Jersey Neuroscience Institute/JFK Medical Center, Edison, NJ, and The Neurogenetics Foundation, Cranbury, NJ. We acknowledge Dr Souvik Sen for his advice and encouragement in the initiation and design of this study.

**MIAMISR and NOMAS:** The Northern Manhattan Study (NOMAS) was supported by grants from the NINDS (R37 NS029993, R01 NS27517). The Cerebrovascular Biorepository at University of Miami/Jackson Memorial Hospital (The Miami Stroke Registry, Institutional Review Board No. 20070386) was supported by the Department of Neurology at University of Miami Miller School of Medicine and Evelyn McKnight Brain Institute. Biorepository and DNA extraction services were provided by the Hussmann Institute for Human Genomics at the Miller School of Medicine.

**MGH-GASROS:** The Massachusetts General Hospital Stroke Genetics Group was supported by the NIH Genes Affecting Stroke Risks and Outcomes Study (GASROS) grant K23 NS042720, the American Heart Association/Bugher Foundation Centers for Stroke Prevention Research 0775010N, and NINDS K23NS042695, K23 NS064052, the Deane Institute for Integrative Research in Atrial Fibrillation and Stroke, and by the Keane Stroke Genetics Fund. Genotyping services were provided by the Broad Institute Center for Genotyping and Analysis, supported by grant U54 RR020278 from the National Center for Research Resources.

**MUNICH:** The MUNICH study was supported by the Vascular Dementia Research Foundation, the Dr. Werner Jackstaedt-Stiftung, the FP7 EU project CVgenes@target (261123), the German Federal Ministry of Education and Research (BMBF) in the context of the e:Med program (e:AtheroSysMed), and by grants from the Deutsche Forschungsgemeinschaft (SFB1123 B3/C1, Munich Cluster for Systems Neurology).

**NHS:** The Nurses’ Health Study work on stroke is supported by grants from the NIH, including HL088521 and HL34594 from the National Heart, Lung, and Blood Institute, as well as grants from the National Cancer Institute funding the questionnaire follow-up and blood collection: CA87969 and CA49449.

**OXVASC:** The Oxford Vascular Study was supported by the Wellcome Trust, Wolfson Foundation, Stroke Association, Medical Research Council, Dunhill Medical Trust, NIH Research (NIHR), and NIHR Oxford Biomedical Research Centre based at Oxford University Hospitals NHS Trust and University of Oxford. Dr Rothwell is in receipt of Senior Investigator Awards from the Wellcome Trust and the NIHR.

**REGARDS:** The Reasons for Geographic and Racial Differences in Stroke (**REGARDS**) Study was supported by a cooperative agreement U01 NS041588 from the NINDS, NIH, and Department of Health and Human Service. A full list of participating REGARDS investigators and institutions can be found at <http://www.regardsstudy.org>.

**SAHLSIS:** The Sahlgrenska Academy Study of Ischemic Stroke was supported by the Swedish Research Council (K2014-64X-14605-12-5), the Swedish Heart and Lung Foundation (20130315), the Swedish state/Sahlgrenska University Hospital (ALFGBG-429981), the Swedish Stroke Association, the Swedish Society of Medicine, and the Rune and Ulla Amlöv Foundation.

**ST. GEORGE’S:** The principal funding for this study was provided by the Wellcome Trust, as part of the Wellcome Trust Case Control Consortium 2 project (085475/B/08/Z and 085475/Z/08/Z and WT084724MA). Collection of some of the St George’s stroke cohort was supported by project grant support from the Stroke Association. Hugh Markus is supported by an NIHR Investigator award. Matthew Traylor is supported by project grant funding from the Stroke Association (TSA 2013/01).

**VISP:** The GWAS component of the VISP study was supported by the United States National Human Genome Research Institute (NHGRI), Grant U01 HG005160 (PI Michèle Sale & Bradford Worrall), as part of the Genomics and Randomized Trials Network (GARNET). Genotyping services were provided by the Johns Hopkins University Center for Inherited Disease Research (CIDR), which is fully funded through a federal contract from the NIH to Johns Hopkins University. Assistance with data cleaning was provided by the GARNET Coordinating Center (U01HG005157; PI Bruce S Weir). Study recruitment and collection of datasets for the VISP clinical trial were supported by an investigator-initiated research grant (R01 NS34447; PI James Toole) from the United States Public Health Service, NINDS, Bethesda, Maryland. Control data for comparison with European ancestry VISP stroke cases were obtained through the database of genotypes and phenotypes (dbGAP) High Density SNP Association Analysis of Melanoma: Case-Control and Outcomes Investigation (phs000187.v1.p1; R01CA100264, 3P50CA093459, 5P50CA097007, 5R01ES011740, 5R01CA133996, HHSN268200782096C; PIs Christopher Amos, Qingyi Wei, Jeffrey E. Lee). For VISP stroke cases of African ancestry, a subset of the Healthy Aging in Neighborhoods of Diversity across the Life Span study (HANDLS) were used as stroke free controls. HANDLS is funded by the National Institute of Aging (1Z01AG000513; PI Michele K. Evans).

**WUSTL:** Washington University St. Louis Stroke Study (**WUSTL**): The collection, extraction of DNA from blood, and storage of specimens were supported by 2 NINDS NIH grants (P50 NS055977 and R01 NS8541901). Basic demographic and clinical characterization of stroke phenotype was prospectively collected in the Cognitive Rehabilitation and Recovery Group (CRRG) registry. The Recovery Genomics after Ischemic Stroke (ReGenesIS) study was supported by a grant from the Barnes-Jewish Hospital Foundation.

### Consortia and study specific affiliations

#### FinnGen Consortium Members

**Aarno Palotie**
Institute for Molecular Medicine Finland (FIMM), HiLIFE, University of Helsinki, Helsinki, Finland; Broad Institute of MIT and Harvard; Massachusetts General Hospital

**Mark Daly**
Institute for Molecular Medicine Finland (FIMM), HiLIFE, University of Helsinki, Helsinki, Finland; Broad Institute of MIT and Harvard; Massachusetts General Hospital

**Bridget Riley-Gills**
Abbvie, Chicago, IL, United States

**Howard Jacob**
Abbvie, Chicago, IL, United States

**Dirk Paul**
Astra Zeneca, Cambridge, United Kingdom

**Slavé Petrovski**
Astra Zeneca, Cambridge, United Kingdom

**Heiko Runz**
Biogen, Cambridge, MA, United States

**Sally John**
Biogen, Cambridge, MA, United States

**George Okafo**
Boehringer Ingelheim, Ingelheim am Rhein, Germany

**Nathan Lawless**
Boehringer Ingelheim, Ingelheim am Rhein, Germany

**Heli Salminen-Mankonen**
Boehringer Ingelheim, Ingelheim am Rhein, Germany

**Robert Plenge**
Bristol Myers Squibb, New York, NY, United States

**Joseph Maranville**
Bristol Myers Squibb, New York, NY, United States

**Mark McCarthy**
Genentech, San Francisco, CA, United States

**Margaret G. Ehm**
GlaxoSmithKline, Collegeville, PA, United States

**Kirsi Auro**
GlaxoSmithKline, Espoo, Finland

**Simonne Longerich**
Merck, Kenilworth, NJ, United States

**Anders Mälarstig**
Pfizer, New York, NY, United States

**Katherine Klinger**
Translational Sciences, Sanofi R&D, Framingham, MA, USA

**Clement Chatelain**
Translational Sciences, Sanofi R&D, Framingham, MA, USA

**Matthias Gossel**
Translational Sciences, Sanofi R&D, Framingham, MA, USA

**Karol Estrada**
Maze Therapeutics, San Francisco, CA, United States

**Robert Graham**
Maze Therapeutics, San Francisco, CA, United States

**Robert Yang**
Janssen Biotech, Beerse, Belgium

**Chris O´Donnell**
Novartis Institutes for BioMedical Research, Cambridge, MA, United States

**Tomi P. Mäkelä**
HiLIFE, University of Helsinki, Finland, Finland

**Jaakko Kaprio**
Institute for Molecular Medicine Finland (FIMM), HiLIFE, University of Helsinki, Helsinki, Finland

**Petri Virolainen**
Auria Biobank / University of Turku / Hospital District of Southwest Finland, Turku, Finland

**Antti Hakanen**
Auria Biobank / University of Turku / Hospital District of Southwest Finland, Turku, Finland

**Terhi Kilpi**
THL Biobank / Finnish Institute for Health and Welfare (THL), Helsinki, Finland

**Markus Perola**
THL Biobank / Finnish Institute for Health and Welfare (THL), Helsinki, Finland

**Jukka Partanen**
Finnish Red Cross Blood Service / Finnish Hematology Registry and Clinical Biobank, Helsinki, Finland

**Anne Pitkäranta**
Helsinki Biobank / Helsinki University and Hospital District of Helsinki and Uusimaa, Helsinki

**Taneli Raivio**
Helsinki Biobank / Helsinki University and Hospital District of Helsinki and Uusimaa, Helsinki

**Jani Tikkanen**
Northern Finland Biobank Borealis / University of Oulu / Northern Ostrobothnia Hospital District, Oulu, Finland

**Raisa Serpi**
Northern Finland Biobank Borealis / University of Oulu / Northern Ostrobothnia Hospital District, Oulu, Finland

**Tarja Laitinen**
Finnish Clinical Biobank Tampere / University of Tampere / Pirkanmaa Hospital District, Tampere, Finland

**Veli-Matti Kosma**
Biobank of Eastern Finland / University of Eastern Finland / Northern Savo Hospital District, Kuopio, Finland

**Jari Laukkanen**
Central Finland Biobank / University of Jyväskylä / Central Finland Health Care District, Jyväskylä, Finland

**Marco Hautalahti**
FINBB - Finnish biobank cooperative

**Outi Tuovila**
Business Finland, Helsinki, Finland

**Raimo Pakkanen**
Business Finland, Helsinki, Finland

**Jeffrey Waring**
Abbvie, Chicago, IL, United States

**Bridget Riley-Gillis**
Abbvie, Chicago, IL, United States

**Fedik Rahimov**
Abbvie, Chicago, IL, United States

**Ioanna Tachmazidou**
Astra Zeneca, Cambridge, United Kingdom

**Chia-Yen Chen**
Biogen, Cambridge, MA, United States

**Zhihao Ding**
Boehringer Ingelheim, Ingelheim am Rhein, Germany

**Marc Jung**
Boehringer Ingelheim, Ingelheim am Rhein, Germany

**Shameek Biswas**
Bristol Myers Squibb, New York, NY, United States

**Rion Pendergrass**
Genentech, San Francisco, CA, United States

**David Pulford**
GlaxoSmithKline, Stevenage, United Kingdom

**Neha Raghavan**
Merck, Kenilworth, NJ, United States

**Adriana Huertas-Vazquez**
Merck, Kenilworth, NJ, United States

**Jae-Hoon Sul**
Merck, Kenilworth, NJ, United States

**Xinli Hu**
Pfizer, New York, NY, United States

**Åsa Hedman**
Pfizer, New York, NY, United States

**Manuel Rivas**
Maze Therapeutics, San Francisco, CA, United States

**Dawn Waterworth**
Janssen Research & Development, LLC, Spring House, PA, United States

**Nicole Renaud**
Novartis Institutes for BioMedical Research, Cambridge, MA, United States

**Ma´en Obeidat**
Novartis Institutes for BioMedical Research, Cambridge, MA, United States

**Samuli Ripatti**
Institute for Molecular Medicine Finland (FIMM), HiLIFE, University of Helsinki, Helsinki, Finland

**Johanna Schleutker**
Auria Biobank / Univ. of Turku / Hospital District of Southwest Finland, Turku, Finland

**Mikko Arvas**
Finnish Red Cross Blood Service / Finnish Hematology Registry and Clinical Biobank, Helsinki, Finland

**Olli Carpén**
Helsinki Biobank / Helsinki University and Hospital District of Helsinki and Uusimaa, Helsinki

**Reetta Hinttala**
Northern Finland Biobank Borealis / University of Oulu / Northern Ostrobothnia Hospital District, Oulu, Finland

**Johannes Kettunen**
Northern Finland Biobank Borealis / University of Oulu / Northern Ostrobothnia Hospital District, Oulu, Finland

**Arto Mannermaa**
Biobank of Eastern Finland / University of Eastern Finland / Northern Savo Hospital District, Kuopio, Finland

**Katriina Aalto-Setälä**
Faculty of Medicine and Health Technology, Tampere University, Tampere, Finland

**Mika Kähönen**
Finnish Clinical Biobank Tampere / University of Tampere / Pirkanmaa Hospital District, Tampere, Finland

**Johanna Mäkelä**
FINBB - Finnish biobank cooperative

**Reetta Kälviäinen**
Northern Savo Hospital District, Kuopio, Finland

**Valtteri Julkunen**
Northern Savo Hospital District, Kuopio, Finland

**Hilkka Soininen**
Northern Savo Hospital District, Kuopio, Finland

**Anne Remes**
Northern Ostrobothnia Hospital District, Oulu, Finland

**Mikko Hiltunen**
University of Eastern Finland, Kuopio, Finland

**Jukka Peltola**
Pirkanmaa Hospital District, Tampere, Finland

**Minna Raivio**
Hospital District of Helsinki and Uusimaa, Helsinki, Finland

**Pentti Tienari**
Hospital District of Helsinki and Uusimaa, Helsinki, Finland

**Juha Rinne**
Hospital District of Southwest Finland, Turku, Finland

**Roosa Kallionpää**
Hospital District of Southwest Finland, Turku, Finland

**Juulia Partanen**
Institute for Molecular Medicine Finland, HiLIFE, University of Helsinki, Finland

**Ali Abbasi**
Abbvie, Chicago, IL, United States

**Adam Ziemann**
Abbvie, Chicago, IL, United States

**Nizar Smaoui**
Abbvie, Chicago, IL, United States

**Anne Lehtonen**
Abbvie, Chicago, IL, United States

**Susan Eaton**
Biogen, Cambridge, MA, United States

**Sanni Lahdenperä**
Biogen, Cambridge, MA, United States

**Natalie Bowers**
Genentech, San Francisco, CA, United States

**Edmond Teng**
Genentech, San Francisco, CA, United States

**Fanli Xu**
GlaxoSmithKline, Brentford, United Kingdom

**Laura Addis**
GlaxoSmithKline, Brentford, United Kingdom

**John Eicher**
GlaxoSmithKline, Brentford, United Kingdom

**Qingqin S Li**
Janssen Research & Development, LLC, Titusville, NJ 08560, United States

**Karen He**
Janssen Research & Development, LLC, Spring House, PA, United States

**Ekaterina Khramtsova**
Janssen Research & Development, LLC, Spring House, PA, United States

**Martti Färkkilä**
Hospital District of Helsinki and Uusimaa, Helsinki, Finland

**Jukka Koskela**
Hospital District of Helsinki and Uusimaa, Helsinki, Finland

**Sampsa Pikkarainen**
Hospital District of Helsinki and Uusimaa, Helsinki, Finland

**Airi Jussila**
Pirkanmaa Hospital District, Tampere, Finland

**Katri Kaukinen**
Pirkanmaa Hospital District, Tampere, Finland

**Timo Blomster**
Northern Ostrobothnia Hospital District, Oulu, Finland

**Mikko Kiviniemi**
Northern Savo Hospital District, Kuopio, Finland

**Markku Voutilainen**
Hospital District of Southwest Finland, Turku, Finland

**Tim Lu**
Genentech, San Francisco, CA, United States

**Linda McCarthy**
GlaxoSmithKline, Brentford, United Kingdom

**Amy Hart**
Janssen Research & Development, LLC, Spring House, PA, United States

**Meijian Guan**
Janssen Research & Development, LLC, Spring House, PA, United States

**Jason Miller**
Merck, Kenilworth, NJ, United States

**Kirsi Kalpala**
Pfizer, New York, NY, United States

**Melissa Miller**
Pfizer, New York, NY, United States

**Kari Eklund**
Hospital District of Helsinki and Uusimaa, Helsinki, Finland

**Antti Palomäki**
Hospital District of Southwest Finland, Turku, Finland

**Pia Isomäki**
Pirkanmaa Hospital District, Tampere, Finland

**Laura Pirilä**
Hospital District of Southwest Finland, Turku, Finland

**Oili Kaipiainen-Seppänen**
Northern Savo Hospital District, Kuopio, Finland

**Johanna Huhtakangas**
Northern Ostrobothnia Hospital District, Oulu, Finland

**Nina Mars**
Institute for Molecular Medicine Finland (FIMM), HiLIFE, University of Helsinki, Helsinki, Finland

**Apinya Lertratanakul**
Abbvie, Chicago, IL, United States

**Coralie Viollet**
AstraZeneca, Cambridge, United Kingdom

**Marla Hochfeld**
Bristol Myers Squibb, New York, NY, United States

**Jorge Esparza Gordillo**
GlaxoSmithKline, Brentford, United Kingdom

**Fabiana Farias**
Merck, Kenilworth, NJ, United States

**Nan Bing**
Pfizer, New York, NY, United States

**Margit Pelkonen**
Northern Savo Hospital District, Kuopio, Finland

**Paula Kauppi**
Hospital District of Helsinki and Uusimaa, Helsinki, Finland

**Hannu Kankaanranta**
University of Gothenburg, Gothenburg, Sweden/ Seinäjoki Central Hospital, Seinäjoki, Finland/ Tampere University, Tampere, Finland

**Terttu Harju**
Northern Ostrobothnia Hospital District, Oulu, Finland

**Riitta Lahesmaa**
Hospital District of Southwest Finland, Turku, Finland

**Hubert Chen**
Genentech, San Francisco, CA, United States

**Joanna Betts**
GlaxoSmithKline, Brentford, United Kingdom

**Rajashree Mishra**
GlaxoSmithKline, Brentford, United Kingdom

**Majd Mouded**
Novartis, Basel, Switzerland

**Debby Ngo**
Novartis, Basel, Switzerland

**Teemu Niiranen**
Finnish Institute for Health and Welfare (THL), Helsinki, Finland

**Felix Vaura**
Finnish Institute for Health and Welfare (THL), Helsinki, Finland

**Veikko Salomaa**
Finnish Institute for Health and Welfare (THL), Helsinki, Finland

**Kaj Metsärinne**
Hospital District of Southwest Finland, Turku, Finland

**Jenni Aittokallio**
Hospital District of Southwest Finland, Turku, Finland

**Jussi Hernesniemi**
Pirkanmaa Hospital District, Tampere, Finland

**Daniel Gordin**
Hospital District of Helsinki and Uusimaa, Helsinki, Finland

**Juha Sinisalo**
Hospital District of Helsinki and Uusimaa, Helsinki, Finland

**Marja-Riitta Taskinen**
Hospital District of Helsinki and Uusimaa, Helsinki, Finland

**Tiinamaija Tuomi**
Hospital District of Helsinki and Uusimaa, Helsinki, Finland

**Timo Hiltunen**
Hospital District of Helsinki and Uusimaa, Helsinki, Finland

**Amanda Elliott**
Institute for Molecular Medicine Finland (FIMM), HiLIFE, University of Helsinki, Helsinki, Finland; Broad Institute, Cambridge, MA, USA and Massachusetts General Hospital, Boston, MA, USA

**Mary Pat Reeve**
Institute for Molecular Medicine Finland (FIMM), HiLIFE, University of Helsinki, Helsinki, Finland

**Sanni Ruotsalainen**
Institute for Molecular Medicine Finland (FIMM), HiLIFE, University of Helsinki, Helsinki, Finland

**Audrey Chu**
GlaxoSmithKline, Brentford, United Kingdom

**Dermot Reilly**
Janssen Research & Development, LLC, Boston, MA, United States

**Mike Mendelson**
Novartis, Boston, MA, United States

**Jaakko Parkkinen**
Pfizer, New York, NY, United States

**Tuomo Meretoja**
Hospital District of Helsinki and Uusimaa, Helsinki, Finland

**Heikki Joensuu**
Hospital District of Helsinki and Uusimaa, Helsinki, Finland

**Johanna Mattson**
Hospital District of Helsinki and Uusimaa, Helsinki, Finland

**Eveliina Salminen**
Hospital District of Helsinki and Uusimaa, Helsinki, Finland

**Annika Auranen**
Pirkanmaa Hospital District , Tampere, Finland

**Peeter Karihtala**
Northern Ostrobothnia Hospital District, Oulu, Finland

**Päivi Auvinen**
Northern Savo Hospital District, Kuopio, Finland

**Klaus Elenius**
Hospital District of Southwest Finland, Turku, Finland

**Esa Pitkänen**
Institute for Molecular Medicine Finland (FIMM), HiLIFE, University of Helsinki, Helsinki, Finland

**Relja Popovic**
Abbvie, Chicago, IL, United States

**Margarete Fabre**
AstraZeneca, Cambridge, United Kingdom

**Jennifer Schutzman**
Genentech, San Francisco, CA, United States

**Diptee Kulkarni**
GlaxoSmithKline, Brentford, United Kingdom

**Alessandro Porello**
Janssen Research & Development, LLC, Spring House, PA, United States

**Andrey Loboda**
Merck, Kenilworth, NJ, United States

**Heli Lehtonen**
Pfizer, New York, NY, United States

**Stefan McDonough**
Pfizer, New York, NY, United States

**Sauli Vuoti**
Janssen-Cilag Oy, Espoo, Finland

**Kai Kaarniranta**
Northern Savo Hospital District, Kuopio, Finland; Department of Molecular Genetics, University of Lodz, Lodz, Poland

**Joni A Turunen**
Helsinki University Hospital and University of Helsinki, Helsinki, Finland; Eye Genetics Group, Folkhälsan Research Center, Helsinki, Finland

**Terhi Ollila**
Hospital District of Helsinki and Uusimaa, Helsinki, Finland

**Hannu Uusitalo**
Pirkanmaa Hospital District, Tampere, Finland

**Juha Karjalainen**
Institute for Molecular Medicine Finland (FIMM), HiLIFE, University of Helsinki, Helsinki, Finland

**Mengzhen Liu**
Abbvie, Chicago, IL, United States

**Stephanie Loomis**
Biogen, Cambridge, MA, United States

**Erich Strauss**
Genentech, San Francisco, CA, United States

**Hao Chen**
Genentech, San Francisco, CA, United States

**Kaisa Tasanen**
Northern Ostrobothnia Hospital District, Oulu, Finland

**Laura Huilaja**
Northern Ostrobothnia Hospital District, Oulu, Finland

**Katariina Hannula-Jouppi**
Hospital District of Helsinki and Uusimaa, Helsinki, Finland

**Teea Salmi**
Pirkanmaa Hospital District, Tampere, Finland

**Sirkku Peltonen**
Hospital District of Southwest Finland, Turku, Finland

**Leena Koulu**
Hospital District of Southwest Finland, Turku, Finland

**David Choy**
Genentech, San Francisco, CA, United States

**Ying Wu**
Pfizer, New York, NY, United States

**Pirkko Pussinen**
Hospital District of Helsinki and Uusimaa, Helsinki, Finland

**Aino Salminen**
Hospital District of Helsinki and Uusimaa, Helsinki, Finland

**Tuula Salo**
Hospital District of Helsinki and Uusimaa, Helsinki, Finland

**David Rice**
Hospital District of Helsinki and Uusimaa, Helsinki, Finland

**Pekka Nieminen**
Hospital District of Helsinki and Uusimaa, Helsinki, Finland

**Ulla Palotie**
Hospital District of Helsinki and Uusimaa, Helsinki, Finland

**Maria Siponen**
Northern Savo Hospital District, Kuopio, Finland

**Liisa Suominen**
Northern Savo Hospital District, Kuopio, Finland

**Päivi Mäntylä**
Northern Savo Hospital District, Kuopio, Finland

**Ulvi Gursoy**
Hospital District of Southwest Finland, Turku, Finland

**Vuokko Anttonen**
Northern Ostrobothnia Hospital District, Oulu, Finland

**Kirsi Sipilä**
Research Unit of Oral Health Sciences Faculty of Medicine, University of Oulu, Oulu, Finland; Medical Research Center, Oulu, Oulu University Hospital and University of Oulu, Oulu, Finland

**Rion Pendergrass**
Genentech, San Francisco, CA, United States

**Hannele Laivuori**
Institute for Molecular Medicine Finland (FIMM), HiLIFE, University of Helsinki, Helsinki, Finland

**Venla Kurra**
Pirkanmaa Hospital District, Tampere, Finland

**Laura Kotaniemi-Talonen**
Pirkanmaa Hospital District, Tampere, Finland

**Oskari Heikinheimo**
Hospital District of Helsinki and Uusimaa, Helsinki, Finland

**Ilkka Kalliala**
Hospital District of Helsinki and Uusimaa, Helsinki, Finland

**Lauri Aaltonen**
Hospital District of Helsinki and Uusimaa, Helsinki, Finland

**Varpu Jokimaa**
Hospital District of Southwest Finland, Turku, Finland

**Marja Vääräsmäki**
Northern Ostrobothnia Hospital District, Oulu, Finland

**Outi Uimari**
Northern Ostrobothnia Hospital District, Oulu, Finland

**Laure Morin-Papunen**
Northern Ostrobothnia Hospital District, Oulu, Finland

**Maarit Niinimäki**
Northern Ostrobothnia Hospital District, Oulu, Finland

**Terhi Piltonen**
Northern Ostrobothnia Hospital District, Oulu, Finland

**Katja Kivinen**
Institute for Molecular Medicine Finland (FIMM), HiLIFE, University of Helsinki, Helsinki, Finland

**Elisabeth Widen**
Institute for Molecular Medicine Finland (FIMM), HiLIFE, University of Helsinki, Helsinki, Finland

**Taru Tukiainen**
Institute for Molecular Medicine Finland (FIMM), HiLIFE, University of Helsinki, Helsinki, Finland

**Niko Välimäki**
University of Helsinki, Helsinki, Finland

**Eija Laakkonen**
University of Jyväskylä, Jyväskylä, Finland

**Jaakko Tyrmi**
University of Oulu, Oulu, Finland / University of Tampere, Tampere, Finland

**Heidi Silven**
University of Oulu, Oulu, Finland

**Eeva Sliz**
University of Oulu, Oulu, Finland

**Riikka Arffman**
University of Oulu, Oulu, Finland

**Susanna Savukoski**
University of Oulu, Oulu, Finland

**Triin Laisk**
Estonian biobank, Tartu, Estonia

**Natalia Pujol**
Estonian biobank, Tartu, Estonia

**Janet Kumar**
GlaxoSmithKline, Collegeville, PA, United States

**Iiris Hovatta**
University of Helsinki, Finland

**Erkki Isometsä**
Hospital District of Helsinki and Uusimaa, Helsinki, Finland

**Hanna Ollila**
Institute for Molecular Medicine Finland (FIMM), HiLIFE, University of Helsinki, Helsinki, Finland

**Jaana Suvisaari**
Finnish Institute for Health and Welfare (THL), Helsinki, Finland

**Thomas Damm Als**
Aarhus University, Denmark

**Antti Mäkitie**
Department of Otorhinolaryngology - Head and Neck Surgery, University of Helsinki and Helsinki University Hospital, Helsinki, Finland

**Argyro Bizaki-Vallaskangas**
Pirkanmaa Hospital District, Tampere, Finland

**Sanna Toppila-Salmi**
University of Helsinki, Finland

**Tytti Willberg**
Hospital District of Southwest Finland, Turku, Finland

**Elmo Saarentaus**
Institute for Molecular Medicine Finland (FIMM), HiLIFE, University of Helsinki, Helsinki, Finland

**Antti Aarnisalo**
Hospital District of Helsinki and Uusimaa, Helsinki, Finland

**Elisa Rahikkala**
Northern Ostrobothnia Hospital District, Oulu, Finland

**Kristiina Aittomäki**
Department of Medical Genetics, Helsinki University Central Hospital, Helsinki, Finland

**Fredrik Åberg**
Transplantation and Liver Surgery Clinic, Helsinki University Hospital, Helsinki University, Helsinki, Finland

**Mitja Kurki**
Institute for Molecular Medicine Finland (FIMM), HiLIFE, University of Helsinki, Helsinki, Finland; Broad Institute, Cambridge, MA, United States

**Aki Havulinna**
Institute for Molecular Medicine Finland (FIMM), HiLIFE, University of Helsinki, Helsinki, Finland; Finnish Institute for Health and Welfare (THL), Helsinki, Finland

**Juha Mehtonen**
Institute for Molecular Medicine Finland (FIMM), HiLIFE, University of Helsinki, Helsinki, Finland

**Priit Palta**
Institute for Molecular Medicine Finland (FIMM), HiLIFE, University of Helsinki, Helsinki, Finland

**Shabbeer Hassan**
Institute for Molecular Medicine Finland (FIMM), HiLIFE, University of Helsinki, Helsinki, Finland

**Pietro Della Briotta Parolo**
Institute for Molecular Medicine Finland (FIMM), HiLIFE, University of Helsinki, Helsinki, Finland

**Wei Zhou**
Broad Institute, Cambridge, MA, United States

**Mutaamba Maasha**
Broad Institute, Cambridge, MA, United States

**Susanna Lemmelä**
Institute for Molecular Medicine Finland (FIMM), HiLIFE, University of Helsinki, Helsinki, Finland

**Aoxing Liu**
Institute for Molecular Medicine Finland (FIMM), HiLIFE, University of Helsinki, Helsinki, Finland

**Arto Lehisto**
Institute for Molecular Medicine Finland (FIMM), HiLIFE, University of Helsinki, Helsinki, Finland

**Andrea Ganna**
Institute for Molecular Medicine Finland (FIMM), HiLIFE, University of Helsinki, Helsinki, Finland

**Vincent Llorens**
Institute for Molecular Medicine Finland (FIMM), HiLIFE, University of Helsinki, Helsinki, Finland

**Henrike Heyne**
Institute for Molecular Medicine Finland (FIMM), HiLIFE, University of Helsinki, Helsinki, Finland

**Joel Rämö**
Institute for Molecular Medicine Finland (FIMM), HiLIFE, University of Helsinki, Helsinki, Finland

**Rodos Rodosthenous**
Institute for Molecular Medicine Finland (FIMM), HiLIFE, University of Helsinki, Helsinki, Finland

**Satu Strausz**
Institute for Molecular Medicine Finland (FIMM), HiLIFE, University of Helsinki, Helsinki, Finland

**Tuula Palotie**
University of Helsinki and Hospital District of Helsinki and Uusimaa, Helsinki, Finland

**Kimmo Palin**
University of Helsinki, Helsinki, Finland

**Javier Garcia-Tabuenca**
University of Tampere, Tampere, Finland

**Harri Siirtola**
University of Tampere, Tampere, Finland

**Tuomo Kiiskinen**
Institute for Molecular Medicine Finland (FIMM), HiLIFE, University of Helsinki, Helsinki, Finland

**Jiwoo Lee**
Institute for Molecular Medicine Finland (FIMM), HiLIFE, University of Helsinki, Helsinki, Finland; Broad Institute, Cambridge, MA, United States

**Kristin Tsuo**
Institute for Molecular Medicine Finland (FIMM), HiLIFE, University of Helsinki, Helsinki, Finland; Broad Institute, Cambridge, MA, United States

**Kati Kristiansson**
THL Biobank / Finnish Institute for Health and Welfare (THL), Helsinki, Finland

**Kati Hyvärinen**
Finnish Red Cross Blood Service, Helsinki, Finland

**Jarmo Ritari**
Finnish Red Cross Blood Service, Helsinki, Finland

**Katri Pylkäs**
University of Oulu, Oulu, Finland

**Minna Karjalainen**
University of Oulu, Oulu, Finland

**Tuomo Mantere**
Northern Finland Biobank Borealis / University of Oulu / Northern Ostrobothnia Hospital District, Oulu, Finland

**Eeva Kangasniemi**
Finnish Clinical Biobank Tampere / University of Tampere / Pirkanmaa Hospital District, Tampere, Finland

**Sami Heikkinen**
University of Eastern Finland, Kuopio, Finland

**Nina Pitkänen**
Auria Biobank / University of Turku / Hospital District of Southwest Finland, Turku, Finland

**Samuel Lessard**
Translational Sciences, Sanofi R&D, Framingham, MA, USA

**Clément Chatelain**
Translational Sciences, Sanofi R&D, Framingham, MA, USA

**Lila Kallio**
Auria Biobank / University of Turku / Hospital District of Southwest Finland, Turku, Finland

**Tiina Wahlfors**
THL Biobank / Finnish Institute for Health and Welfare (THL), Helsinki, Finland

**Eero Punkka**
Helsinki Biobank / Helsinki University and Hospital District of Helsinki and Uusimaa, Helsinki

**Sanna Siltanen**
Finnish Clinical Biobank Tampere / University of Tampere / Pirkanmaa Hospital District, Tampere, Finland

**Teijo Kuopio**
Central Finland Biobank / University of Jyväskylä / Central Finland Health Care District, Jyväskylä, Finland

**Anu Jalanko**
Institute for Molecular Medicine Finland (FIMM), HiLIFE, University of Helsinki, Helsinki, Finland

**Huei-Yi Shen**
Institute for Molecular Medicine Finland (FIMM), HiLIFE, University of Helsinki, Helsinki, Finland

**Risto Kajanne**
Institute for Molecular Medicine Finland (FIMM), HiLIFE, University of Helsinki, Helsinki, Finland

**Mervi Aavikko**
Institute for Molecular Medicine Finland (FIMM), HiLIFE, University of Helsinki, Helsinki, Finland

**Helen Cooper**
Institute for Molecular Medicine Finland (FIMM), HiLIFE, University of Helsinki, Helsinki, Finland

**Denise Öller**
Institute for Molecular Medicine Finland (FIMM), HiLIFE, University of Helsinki, Helsinki, Finland

**Rasko Leinonen**
Institute for Molecular Medicine Finland (FIMM), HiLIFE, University of Helsinki, Helsinki, Finland; European Molecular Biology Laboratory, European Bioinformatics Institute, Cambridge, UK

**Henna Palin**
Finnish Clinical Biobank Tampere / University of Tampere / Pirkanmaa Hospital District, Tampere, Finland

**Malla-Maria Linna**
Helsinki Biobank / Helsinki University and Hospital District of Helsinki and Uusimaa, Helsinki

**Masahiro Kanai**
Broad Institute, Cambridge, MA, United States

**Zhili Zheng**
Broad Institute, Cambridge, MA, United States

**L. Elisa Lahtela**
Institute for Molecular Medicine Finland (FIMM), HiLIFE, University of Helsinki, Helsinki, Finland

**Mari Kaunisto**
Institute for Molecular Medicine Finland (FIMM), HiLIFE, University of Helsinki, Helsinki, Finland

**Elina Kilpeläinen**
Institute for Molecular Medicine Finland (FIMM), HiLIFE, University of Helsinki, Helsinki, Finland

**Timo P. Sipilä**
Institute for Molecular Medicine Finland (FIMM), HiLIFE, University of Helsinki, Helsinki, Finland

**Oluwaseun Alexander Dada**
Institute for Molecular Medicine Finland (FIMM), HiLIFE, University of Helsinki, Helsinki, Finland

**Awaisa Ghazal**
Institute for Molecular Medicine Finland (FIMM), HiLIFE, University of Helsinki, Helsinki, Finland

**Anastasia Kytölä**
Institute for Molecular Medicine Finland (FIMM), HiLIFE, University of Helsinki, Helsinki, Finland

**Rigbe Weldatsadik**
Institute for Molecular Medicine Finland (FIMM), HiLIFE, University of Helsinki, Helsinki, Finland

**Kati Donner**
Institute for Molecular Medicine Finland (FIMM), HiLIFE, University of Helsinki, Helsinki, Finland

**Anu Loukola**
Helsinki Biobank / Helsinki University and Hospital District of Helsinki and Uusimaa, Helsinki

**Päivi Laiho**
THL Biobank / Finnish Institute for Health and Welfare (THL), Helsinki, Finland

**Tuuli Sistonen**
THL Biobank / Finnish Institute for Health and Welfare (THL), Helsinki, Finland

**Essi Kaiharju**
THL Biobank / Finnish Institute for Health and Welfare (THL), Helsinki, Finland

**Markku Laukkanen**
THL Biobank / Finnish Institute for Health and Welfare (THL), Helsinki, Finland

**Elina Järvensivu**
THL Biobank / Finnish Institute for Health and Welfare (THL), Helsinki, Finland

**Sini Lähteenmäki**
THL Biobank / Finnish Institute for Health and Welfare (THL), Helsinki, Finland

**Lotta Männikkö**
THL Biobank / Finnish Institute for Health and Welfare (THL), Helsinki, Finland

**Regis Wong**
THL Biobank / Finnish Institute for Health and Welfare (THL), Helsinki, Finland

**Auli Toivola**
THL Biobank / Finnish Institute for Health and Welfare (THL), Helsinki, Finland

**Minna Brunfeldt**
THL Biobank / Finnish Institute for Health and Welfare (THL), Helsinki, Finland

**Hannele Mattsson**
THL Biobank / Finnish Institute for Health and Welfare (THL), Helsinki, Finland

**Sami Koskelainen**
THL Biobank / Finnish Institute for Health and Welfare (THL), Helsinki, Finland

**Tero Hiekkalinna**
THL Biobank / Finnish Institute for Health and Welfare (THL), Helsinki, Finland

**Teemu Paajanen**
THL Biobank / Finnish Institute for Health and Welfare (THL), Helsinki, Finland

**Kalle Pärn**
Institute for Molecular Medicine Finland (FIMM), HiLIFE, University of Helsinki, Helsinki, Finland

**Mart Kals**
Institute for Molecular Medicine Finland (FIMM), HiLIFE, University of Helsinki, Helsinki, Finland

**Shuang Luo**
Institute for Molecular Medicine Finland (FIMM), HiLIFE, University of Helsinki, Helsinki, Finland

**Shanmukha Sampath Padmanabhuni**
Institute for Molecular Medicine Finland (FIMM), HiLIFE, University of Helsinki, Helsinki, Finland

**Marianna Niemi**
University of Tampere, Tampere, Finland

**Javier Gracia-Tabuenca**
University of Tampere, Tampere, Finland

**Mika Helminen**
University of Tampere, Tampere, Finland

**Tiina Luukkaala**
University of Tampere, Tampere, Finland

**Iida Vähätalo**
University of Tampere, Tampere, Finland

**Jyrki Tammerluoto**
Institute for Molecular Medicine Finland (FIMM), HiLIFE, University of Helsinki, Helsinki, Finland

**Sarah Smith**
Finnish Biobank Cooperative - FINBB

**Tom Southerington**
Finnish Biobank Cooperative - FINBB

**Petri Lehto**
Finnish Biobank Cooperative - FINBB

#### SiGN Author Names and Affiliations

**Sara L. Pulit, PhD**

Department of Medical Genetics, Institute for Molecular Medicine, University Medical Center Utrecht, Utrecht, The Netherlands

**Patrick F. McArdle, PhD**

Department of Medicine and Program for Personalized and Genomic Medicine, University of Maryland School of Medicine, Baltimore, MD

**Quenna Wong, MS**

Department of Biostatistics, University of Washington, Seattle, WA, USA

**Rainer Malik, PhD**

Institute for Stroke and Dementia Research, Klinikum der Universität München, Ludwig-Maximilians University, Munich, Germany

**Katrina Gwinn, MD**

National Institute of Neurological Disorders and Stroke, National Institutes of Health, Bethesda, MD, USA

**Sefanja Achterberg, MD**

Department of Neurology and Neurosurgery, Brain Center Rudolf Magnus, University Medical Center Utrecht, Utrecht, The Netherlands

**Ale Algra, MD**

Department of Neurology and Neurosurgery, Brain Center Rudolf Magnus, University Medical Center Utrecht, Utrecht, The Netherlands

Julius Center for Health Sciences and Primary Care, University Medical Center Utrecht, Utrecht, The Netherlands

**Philippe Amouyel, MD, PhD**
INSERM U-1167, Lille, France Institut Pasteur de Lille, Lille, France

Université Lille Nord de France, Lille, France Lille University Hospital, Lille, France

**Christopher D. Anderson, MD, MMSc**

Department of Neurology and Center for Human Genetic Research Massachusetts General Hospital, Boston, MA, USA

Program in Medical and Population Genetics

Broad Institute of MIT and Harvard, Cambridge, MA, USA Harvard Medical School, Boston, MA, USA

**Donna K. Arnett, PhD, MSPH**

Department of Epidemiology, School of Public Health, University of Alabama at Birmingham, Birmingham, AL, USA

**Ethem Murat Arsava, MD**

AA Martinos Center for Biomedical Imaging, Department of Radiology, Massachusetts General Hospital, Harvard Medical School, Boston, MA, USA

**John Attia, MD, PhD, FRACP, FRCPC**

School of Medicine and Public Health, University of Newcastle, NSW, Australia Division of General Medicine, John Hunter Hospital, Newcastle, NSW, Australia

**Hakan Ay, MD**

AA Martinos Center for Biomedical Imaging, Department of Radiology, Massachusetts General Hospital, Harvard Medical School, Boston, MA, USA Stroke Service, Department of Neurology, Massachusetts General Hospital, Harvard Medical School, Boston, MA, USA

**Traci M. Bartz, MS**

Department of Biostatistics, University of Washington, Seattle, WA, USA

**Thomas Battey, BA**

Department of Neurology and Center for Human Genetic Research, Massachusetts General Hospital, Boston, MA, USA

**Oscar R. Benavente, MD, FRCP**

Department of Neurology, University of British Columbia, Vancouver, British Columbia, Canada

**Steve Bevan, PhD**

Department of Clinical Neurosciences, University of Cambridge, Cambridge, UK

**Alessandro Biffi, MD**

Department of Neurology and Center for Human Genetic Research, Massachusetts General Hospital, Boston, MA, USA

**Joshua C. Bis, PhD**

Cardiovascular Health Research Unit, Department of Medicine, University of Washington, Seattle, WA, USA

**Susan H. Blanton, PhD**

John P. Hussman Institute for Human Genomics, Miller School of Medicine, University of Miami, Miami, FL, USA

**Giorgio B. Boncoraglio MD**

Department of Cerebrovascular Diseases,

Fondazione IRCCS Istituto Neurologico Carlo Besta, Milano, Italy

**Robert D. Brown, Jr., MD, MPH**

Department of Neurology, Mayo Clinic, Rochester, MN, USA

**Annette I. Burgess DPhil**

Stroke Prevention Research Unit, Nuffield Department of Clinical Neurosciences, University of Oxford, John Radcliffe Hospital, Oxford, UK

**Caty Carrera MD MSc**

Neurovascular Research Laboratory. Vall d’Hebron Institute of Research, Vall d’Hebron Hospital, Universitat Autonoma Barcelona, Barcelona, Spain

**Sherita N. Chapman Smith, MD**

Department of Neurology, Virginia Commonwealth University, Richmond, VA, USA

**Daniel I. Chasman, PhD**

Division of Preventive Medicine, Brigham and Women's Hospital, Boston, MA, USA Harvard Medical School, Boston, MA, USA

**Ganesh Chauhan, PhD**

INSERM U897 Neuroepidemiology, Bordeaux, France University of Bordeaux, Bordeaux, France

**Wei-Min Chen, PhD**

Center for Public Health Genomics; Department of Public Health Sciences; and Department of Biochemistry and Molecular Genetics, University of Virginia, Charlottesville, VA, USA

**Yu-Ching Cheng, PhD**

Department of Medicine, University of Maryland School of Medicine, Baltimore, MD, USA

**Michael Chong, MSc**

Population Health Research Institute, McMaster University, DBCVS Research Institute, Hamilton, Ontario, Canada

**Lisa K. Cloonan, BA**

Stroke Division, Department of Neurology, Massachusetts General Hospital, Harvard Medical School, Boston, MA, USA

**John W. Cole, MD, MS**

Department of Neurology, University of Maryland School of Medicine and Veterans Affairs Maryland Health Care System, Baltimore, MD, USA

**Ioana Cotlarciuc, MSc, PhD**

Institute of Cardiovascular Research, Royal Holloway University of London (ICR2UL), Egham, UK

**Carlos Cruchaga, PhD**

Department of Psychiatry, Washington University School of Medicine, St. Louis, MO, USA

**Elisa Cuadrado-Godia, MD**

Department of Neurology, Neurovascular Research Group (NEUVAS)

IMIM-Hospital del Mar (Institut Hospital del Mar d’Investigacions Mèdiques), Universitat Autonoma de Barcelona/DCEXS-Universitat Pompeu Fabra, Barcelona, Spain

**Tushar Dave, MS**

Department of Medicine and Program for Personalized and Genomic Medicine, University of Maryland School of Medicine, Baltimore, MD, USA

**Jesse Dawson, MD, MBChB (hons), FRCP, BSc (hons)**

Institute of Cardiovascular and Medical Sciences, College of Medical, Veterinary & Life Sciences, University of Glasgow, UK

**Stéphanie Debette, MD, PhD**

INSERM U897 Neuroepidemiology, Bordeaux, France University of Bordeaux, Bordeaux, France

Department of Neurology, Bordeaux University Hospital, Bordeaux, France Boston University School of Medicine, Framingham Heart Study, Boston, MA, USA

**Hossein Delavaran, MD**

Department of Clinical Sciences Lund, Neurology, Lund University, Lund, Sweden Department of Neurology and Rehabilitation Medicine, Neurology, Skåne University Hospital, Lund, Sweden

**Cameron A. Dell, BS**

Department of Neurology, University of Maryland School of Medicine, Baltimore, MD, USA

**Martin Dichgans, MD**

Institute for Stroke and Dementia Research, Klinikum der Universität München, Ludwig-Maximilians University, Munich, Germany

Munich Cluster for Systems Neurology (SyNergy), Munich, Germany

**Kimberly F. Doheny, PhD**

Center for Inherited Disease Research, Institute of Genetic Medicine, Johns Hopkins School of Medicine, Baltimore, MD

**Chuanhui Dong, PhD**

Department of Neurology, Miller School of Medicine, University of Miami, Miami, FL, USA

**David J. Duggan, PhD**

Genetic Basis of Human Disease Division, Translational Genomics Research Institute (TGen), Phoenix, AZ, USA

**Gunnar Engström, PhD**

Cardio-vascular Epidemiology, Department of Clinical Sciences Malmö, Lund University, Skåne University Hospital Malmö, Sweden

**Michele K. Evans, MD**

Health Disparities Unit, National Institute on Aging, National Institutes of Health, Baltimore, MD, USA

**Xavier Estivill Pallejà, MD, PhD**

Genomics and Disease group, Center for Genomic Regulation, Barcelona, Spain Universitat Pompeu Fabra (UPF), Barcelona, Spain

Centro de Investigación Biomédica en Red (CIBERESP), Spain

IMIM (Hospital del Mar Medical Research Institute), Barcelona, Spain

**Jessica D. Faul, PhD, MPH**

Survey Research Center, University of Michigan, Ann Arbor, MI, USA

**Israel Fernández-Cadenas, PhD**

Stroke Pharmacogenomics and Genetics, Fundació Docència i Recerca MutuaTerrassa, Mutua de Terrassa Hospital,

Terrassa (Barcelona), Spain

Neurovascular Research Laboratory. Vall d’Hebron Institute of Research, Vall d’Hebron Hospital, Universitat Autonoma Barcelona,

Barcelona, Spain

**Myriam Fornage, PhD**

Institute of Molecular Medicine, University of Texas Health Science Center at Houston, Houston, TX, USA

**Philippe M. Frossard, PhD, DSc**

Center for Non-Communicable Diseases, Karachi, Pakistan Nazarbayev University, Astana, Kazakhstan

**Karen Furie, MD, MPH**

Department of Neurology, Warren Alpert Medical School of Brown University, Providence, RI, USA

**Dale M. Gamble, MHSc, CCRP**

Department of Neurology, Mayo Clinic, Jacksonville, FL, USA

**Christian Gieger, PhD**

Research unit of Molecular Epidemiology, Helmholtz Zentrum München - German Research Center for Environmental Health, Neuherberg, Germany

Institute of Epidemiology II, Helmholtz Zentrum München - German Research Center for Environmental Health, Neuherberg, Germany.

**Anne-Katrin Giese, MD**

Albrecht-Kossel-Institute for Neuroregeneration, Medical University of Rostock, Rostock, Germany

**Eva Giralt-Steinhauer, MD**

Department of Neurology, Neurovascular Research Group (NEUVAS)

IMIM-Hospital del Mar (Institut Hospital del Mar d’Investigacions Mèdiques), Barcelona, Spain

**Hector M. González, PhD**

Department of Epidemiology & Biostatistics, Michigan State University, East Lansing, MI, USA

**An Goris, PhD**

Department of Neurosciences, Laboratory for Neuroimmunology, KU Leuven- University of Leuven, Leuven, Belgium

**Solveig Gretarsdottir, PhD**

deCODE Genetics/Amgen, Reykjavik, Iceland

**Raji P. Grewal, MD**

Neuroscience Institute, Saint Francis Medical Center, School of Health and Medical Sciences, Seton Hall University, South Orange, New Jersey, USA

**Ulrike Grittner, PhD**

Department for Biostatistics and Clinical Epidemiology. Charité-University Medical Centre, Berlin, Germany

**Stefan Gustafsson, PhD, MSc**

Department of Medical Sciences, Molecular Epidemiology and Science for Life Laboratory, Uppsala University, Uppsala, Sweden.

**Buhm Han, PhD**

Asan Institute for Life Sciences, Asan Medical Center, Seoul 138-736, Republic of Korea

Department of Medicine, University of Ulsan College of Medicine, Seoul 138-736, Republic of Korea

**Graeme J. Hankey, MD, FRACP, FRCP, FAHA**

School of Medicine and Pharmacology, The University of Western Australia, Perth, Australia

**Laura Heitsch, MD**

Division of Emergency Medicine, Department of Internal Medicine, Washington University School of Medicine, St. Louis, MO, USA

**Peter Higgins, MD**

Institute of Cardiovascular and Medical Sciences, University of Glasgow, Glasgow, UK

**Marc C. Hochberg, MD**

Division of Rheumatology and Clinical Immunology, Department of Medicine and Department of Epidemiology and Public Health, University of Maryland School of Medicine, Baltimore, MD

**Elizabeth Holliday, BSc(Hons), MSc, PhD**

Public Health Research Program, Hunter Medical Research Institute, Newcastle, NSW, Australia

**Jemma C. Hopewell, PhD**

Clinical Trial Service Unit and Epidemiological Studies Unit, University of Oxford, UK

**Richard B. Horenstein, MD, JD**

Division of Endocrinology, Diabetes and Nutrition, University of Maryland School of Medicine, Baltimore, MD, USA

Program for Personalized and Genomic Medicine, University of Maryland School of Medicine, Baltimore, MD, USA

**George Howard, DrPH**

Department of Biostatistics, School of Public Health, University of Alabama at Birmingham, Birmingham, AL, USA

**M. Arfan Ikram, MD, PhD**

Department of Epidemiology, Erasmus MC University Medical Center, Rotterdam, The Netherlands

**Andreea Ilinca MD**

Department of Clinical Sciences Lund, Neurology, Lund University, Lund, Sweden Department of Internal Medicine, Neurology, Landskrona Hospital, Sweden

**Erik Ingelsson, MD, PhD, FAHA**

Department of Medical Sciences, Molecular Epidemiology and Science for Life Laboratory, Uppsala University, Uppsala, Sweden.

Wellcome Trust Centre for Human Genetics, University of Oxford, Oxford, OX3 7BN, United Kingdom.

**Marguerite R. Irvin, PhD**

Department of Epidemiology, School of Public Health, University of Alabama at Birmingham, Birmingham, AL, USA

**Rebecca D. Jackson, MD**

Division of Endocrinology, Diabetes and Metabolism, Department of Internal Medicine and the Center for Clinical and Translational Science, The Ohio State University, Columbus, OH.

**Christina Jern, MD, PhD**

Institute of Biomedicine, the Sahlgrenska Academy at University of Gothenburg, Gothenburg, Sweden

**Jordi Jiménez Conde, MD, PhD**

Department of Neurology, Neurovascular Research Group (NEUVAS)

IMIM-Hospital del Mar (Institut Hospital del Mar d’Investigacions Mèdiques), Universitat Autonoma de Barcelona/DCEXS-Universitat Pompeu Fabra, Barcelona, Spain

**Julie A. Johnson, PharmD**

Department of Pharmacotherapy and Translational Research and Center for Pharmacogenomics, College of Pharmacy, University of Florida, Gainesville FL, USA

Division of Cardiovascular Medicine, College of Medicine, University of Florida, Gainesville, FL, USA

**Katarina Jood, MD, PhD**

Institute of Neuroscience, the Sahlgrenska Academy at University of Gothenburg, Gothenburg, Sweden

**Muhammed S. Kahn, MSc**

Institute of Cardiovascular Research, Royal Holloway University of London (ICR2UL), Egham, UK

St Peter’s and Ashford Hospitals, UK

**Robert Kaplan, PhD**

Department of Epidemiology and Population Health, Albert Einstein College of Medicine, Bronx, NY, USA

**L. Jaap Kappelle, MD**

Department of Neurology and Neurosurgery, Brain Center Rudolf Magnus, University Medical Center Utrecht, Utrecht, The Netherlands

**Sharon LR Kardia, PhD**

Department of Epidemiology, School of Public Health, University of Michigan, Ann Arbor, MI, USA

**Keith L. Keene PhD**

Department of Biology; Center for Health Disparities, East Carolina University, Greenville, NC, USA

**Brett M. Kissela, MD, MS**

University of Cincinnati College of Medicine, Cincinnati, OH, USA

**Dawn O. Kleindorfer, MD, MS**

University of Cincinnati College of Medicine, Cincinnati, OH, USA

**Simon Koblar, BMBS, FRACP, PhD**

School of Medicine, The Queen Elizabeth Hospital campus, Woodville South SA, Australia

**Daniel Labovitz, MD, MS**

Albert Einstein College of Medicine, Montefiore Medical Center, Bronx, NY, USA

**Lenore J. Launer, PhD**

National Institute on Aging, National Institutes of Health, Bethesda, MD, USA

**Cathy C. Laurie, PhD**

Department of Biostatistics, University of Washington, Seattle, WA, USA

**Cecelia A. Laurie, PhD**

Department of Biostatistics, University of Washington, Seattle, WA, USA

**Cue Hyunkyu Lee, MS**

Asan Institute for Life Sciences, Asan Medical Center, Seoul, Republic of Korea

**Jin-Moo Lee, MD, PhD**

Stroke Center, Department of Neurology, Washington University School of Medicine, St. Louis, MO, USA

**Manuel Lehm**

Institute for Stroke and Dementia Research, Klinikum der Universität München, Ludwig-Maximilians University, Munich, Germany

**Robin Lemmens, MD, PhD**

KU Leuven - University of Leuven, Department of Neurosciences, Experimental Neurology and Leuven Research Institute for Neuroscience and Disease (LIND), Leuven, Belgium

VIB, Vesalius Research Center, Laboratory of Neurobiology, B-3000 Leuven, Belgium

University Hospitals Leuven, Department of Neurology, Leuven, Belgium

**Christopher Levi, MBBS, BMed Sci, FRACP**

John Hunter Hospital, Hunter Medical Research Institute and University of Newcastle, NSW, Australia

**Didier Leys, MD, PhD**

Université Lille Nord de France, Lille, France Lille University Hospital, Lille, France

Department of Neurology, Equipe d’accueil 1046, Lille, France INSERM U897, University of Bordeaux, Bordeaux, France

**Arne Lindgren MD PhD**

Department of Clinical Sciences Lund, Neurology, Lund University, Lund, Sweden Department of Neurology and Rehabilitation Medicine, Neurology, Skåne University Hospital, Lund, Sweden

**W. T. Longstreth Jr., MD**

Department of Neurology, University of Washington, Seattle, WA Department of Epidemiology, University of Washington, Seattle, WA

**Jane Maguire, PhD, BNurs(Hons), BA, RN**

School of Nursing and Midwifery, University of Newcastle, Callaghan, NSW, Australia

**Ani Manichaikul, PhD**

Center for Public Health Genomics, Biostatistics Section, Department for Public Health Sciences, University of Virginia, Charlottesville, VA, USA

**Hugh S. Markus, DM**

Department of Clinical Neurosciences, University of Cambridge, Cambridge, UK

**Leslie A. McClure, PhD**

Department of Biostatistics, School of Public Health, University of Alabama at Birmingham, Birmingham, AL, USA

**Caitrin W. McDonough, PhD**

Department of Pharmacotherapy and Translational Research and Center for Pharmacogenomics, College of Pharmacy, University of Florida, Gainesville FL, USA

**Christa Meisinger, MD**

Institute of Epidemiology II, Helmholtz Zentrum München - German Research Center for Environmental Health, Neuherberg, Germany

**Olle Melander, MD, PhD**

Lund University, Department of Clinical Sciences, Malmö University Hospital, Malmö, Sweden

**James F. Meschia, MD**

Department of Neurology, Mayo Clinic, Jacksonville, FL, USA

**Marina Mola-Caminal, BSc**

Department of Neurology, Neurovascular Research Group (NEUVAS)

IMIM-Hospital del Mar (Institut Hospital del Mar d’Investigacions Mèdiques), Barcelona, Spain

**Joan Montaner, MD, PhD**

Neurovascular Research Laboratory, and Neurology Department

Vall d’Hebron Institute of Research (VHIR), Vall d’Hebron University Hospital, Autonomous University of Barcelona, Barcelona, Spain

**Thomas H. Mosley, PhD**

Department of Medicine (Geriatrics) and Neurology, University of Mississippi Medical Center, Jackson, MS, USA

**Martina Müller-Nurasyid, PhD**

Institute of Genetic Epidemiology, Helmholtz Zentrum München - German Research Center for Environmental Health, Neuherberg, Germany

Department of Medicine I, Ludwig-Maximilians-University Munich, Munich, Germany

DZHK (German Centre for Cardiovascular Research), partner site Munich Heart Alliance, Munich, Germany

**Mike A. Nalls, PhD**

Laboratory of Neurogenetics, National Institute on Aging, National Institutes of Health, Bethesda, MD, USA

**Jeffrey R. O’Connell, DPhil**

Division of Endocrinology, Diabetes and Nutrition, University of Maryland School of Medicine, Baltimore, MD, USA

**Martin O’Donnell, MD, PhD, MRCPI, BCh,BAO**

DBCVS Research Institute, Population Health Research Institute, McMaster University, Hamilton, Ontario, Canada

**Ángel Ois, MD. PhD**

Department of Neurology, Neurovascular Research Group (NEUVAS)

IMIM-Hospital del Mar (Institut Hospital del Mar d’Investigacions Mèdiques), Universitat Autonoma de Barcelona/DCEXS-Universitat Pompeu Fabra, Barcelona, Spain

**George J. Papanicolaou, PhD**

Division of Cardiovascular Sciences, National Heart, Lung, and Blood Institute, Bethesda, MD, USA

**Guillaume Paré, MD, MSc, FRCPC**

DBCVS Research Institute, Population Health Research Institute, McMaster University, Hamilton, Ontario, Canada

**Leema Reddy Peddareddygari, MBBS, MD**

Neuroscience Institute, Saint Francis Medical Center, School of Health and Medical Sciences, Seton Hall University, South Orange, New Jersey, USA

**Annie Pedersén, MD**

Institute of Biomedicine, the Sahlgrenska Academy at University of Gothenburg, Gothenburg, Sweden

**Joanna Pera, MD, PhD**

Department of Neurology, Jagiellonian University Medical College, Krakow, Poland

**Annette Peters, PhD**

Institute of Epidemiology II, Helmholtz Zentrum München - German Research Center for Environmental Health, Neuherberg, Germany

DZHK (German Centre for Cardiovascular Research), partner site Munich Heart Alliance, Munich, Germany

**Deborah Poole, HNC**

Stroke Prevention Research Unit, Nuffield Department of Clinical Neurosciences, University of Oxford, John Radcliffe Hospital, Oxford, UK

**Bruce M. Psaty, MD, PhD**

Cardiovascular Health Research Unit, Department of Medicine, University of Washington, Seattle, WA, USA

Department of Epidemiology, University of Washington, Seattle, WA, USA Department of Health Services, University of Washington, Seattle, WA, USA Group Health Research Institute, Group Health, Seattle, WA, USA

**Raquel Rabionet, PhD**

Genomics and Disease group, Center for Genomic Regulation, Barcelona, Spain Universitat Pompeu Fabra (UPF), Barcelona, Spain

Centro de Investigación Biomédica en Red (CIBERESP), Spain

IMIM (Hospital del Mar Medical Research Institute), Barcelona, Spain

**Miriam R. Raffeld, BA**

Department of Neurology and Center for Human Genetic Research, Massachusetts General Hospital, Boston, MA, USA

**Kristiina Rannikmäe, MD**

Centre for Clinical Brain Sciences, University of Edinburgh, UK

**Asif Rasheed, MBBS**

Center for Non-Communicable Diseases, Karachi, Pakistan

**Petra Redfors, MD, PhD**

Institute of Neuroscience, the Sahlgrenska Academy at University of Gothenburg, Gothenburg, Sweden

**Alex P. Reiner, MD, MSc**

Division of Public Health Sciences, Fred Hutchinson Cancer Research Center, Seattle, WA, USA

**Kathryn Rexrode, MD, MPH**

Brigham and Women's Hospital, Boston, MA, USA

**Marta Ribasés, PhD, BSc**

Psychiatric Genetics Unit, Group of Psychiatry, Mental Health and Addictions, Vall d’Hebron Research Institute (VHIR), Universitat Autònoma de Barcelona, Barcelona, Spain

Department of Psychiatry, Hospital Universitari Vall d’Hebron, Barcelona, Spain Biomedical Network Research Centre on Mental Health (CIBERSAM), Barcelona, Spain

**Stephen S. Rich, PhD**

Center for Public Health Genomics, University of Virginia, Charlottesville, VA USA

**Wim Robberecht, MD, PhD**

KU Leuven - University of Leuven, Department of Neurosciences, Experimental Neurology and Leuven Research Institute for Neuroscience and Disease (LIND), Leuven, Belgium

VIB, Vesalius Research Center, Laboratory of Neurobiology, Leuven, Belgium University Hospitals Leuven, Department of Neurology, Leuven, Belgium

**Ana Rodríguez-Campello, MD**

Department of Neurology, Neurovascular Research Group (NEUVAS)

IMIM-Hospital del Mar (Institut Hospital del Mar d’Investigacions Mèdiques), Universitat Autonoma de Barcelona/DCEXS-Universitat Pompeu Fabra, Barcelona, Spain

**Arndt Rolfs, MD**

Albrecht-Kossel-Institute for Neuroregenation Medical Faculty, University of Rostock, Germany

**Jaume Roquer, MD, PhD**

Department of Neurology, Neurovascular Research Group (NEUVAS)

IMIM-Hospital del Mar (Institut Hospital del Mar d’Investigacions Mèdiques), Universitat Autonoma de Barcelona/DCEXS-Universitat Pompeu Fabra, Barcelona, Spain

**Lynda M. Rose, MS**

Brigham and Women's Hospital, Boston, MA, USA

**Daniel Rosenbaum, MD**

State University of New York, Downstate, Brooklyn, NY, USA

**Natalia S. Rost, MD, MPH**

Stroke Division, Department of Neurology, Massachusetts General Hospital, Harvard Medical School, Boston, MA 02114

**Peter M. Rothwell, FMedSci**

Stroke Prevention Research Unit, Nuffield Department of Clinical Neurosciences, University of Oxford, John Radcliffe Hospital, Oxford, UK

**Tatjana Rundek, MD, PhD, FANA**

Department of Neurology, Miller School of Medicine, University of Miami, Miami, FL, USA

**Kathleen A. Ryan, MPH**

Department of Medicine, University of Maryland School of Medicine, Baltimore, MD, USA

**Ralph L. Sacco, MD, MS, FAHA, FAAN, FANA**

Department of Neurology, Miller School of Medicine, University of Miami, Miami, FL, USA

**Michèle M. Sale, PhD**

Center for Public Health Genomics; Department of Public Health Sciences; and Department of Biochemistry and Molecular Genetics, University of Virginia, VA, USA

**Danish Saleheen, MBBS, PhD**

Department of Biostatistics and Epidemiology, University of Pennsylvania, Philadelphia, PA, USA

Center for Non-Communicable Diseases, Karachi, Pakistan

**Veikko Salomaa, MD, PhD**

National Institute for Health and Welfare, Helsinki, Finland.

**Cristina Sánchez-Mora, PhD, BSc**

Psychiatric Genetics Unit, Group of Psychiatry, Mental Health and Addictions, Vall d’Hebron Research Institute (VHIR), Universitat Autònoma de Barcelona, Barcelona, Spain

Department of Psychiatry, Hospital Universitari Vall d’Hebron, Barcelona, Spain Biomedical Network Research Centre on Mental Health (CIBERSAM), Barcelona, Spain

**Carsten Oliver Schmidt, PD Dr.**

Institute for Community Medicine, University Medicine Greifswald, Greifswald, Germany

**Helena Schmidt, MD, PhD**

Institute of Molecular Biology and Biochemistry, Graz, Austria

**Reinhold Schmidt, MD**

Department of Neurology, Clinical Division of Neurogeriatrics, Medical University Graz, Graz, Austria

**Markus Schürks, MD, MSc**

Department of Neurology, University Hospital Essen, Essen, Germany

**Rodney Scott, BSc(Hons), PhD, FRCPath, FHGSA, FFSc(RCPA)**

School of Biomedical Sciences and Pharmacy, University of Newcastle, NSW, Australia

**Helen C. Segal, PhD**

Stroke Prevention Research Unit, Nuffield Department of Clinical Neurosciences, University of Oxford, John Radcliffe Hospital, Oxford, UK

**Stephan Seiler, MD**

Department of Neurology, Clinical Division of Neurogeriatrics, Medical University Graz, Austria

**Sudha Seshadri, MD**

Department of Neurology, Boston University School of Medicine, Boston, MA, USA

**Pankaj Sharma, MD, PhD, FRCP**

Institute of Cardiovascular Research, Royal Holloway University of London (ICR2UL), Egham, UK

St Peter’s and Ashford Hospitals, UK

**Alan R. Shuldiner, MD**

Division of Endocrinology, Diabetes and Nutrition, University of Maryland School of Medicine, Baltimore, MD, USA

Geriatric Research and Education Clinical Center, Veterans Administration Medical Center, Baltimore, MD, USA

Program for Personalized and Genomic Medicine, University of Maryland School of Medicine, Baltimore, MD, USA

**Brian Silver, MD**

Department of Neurology, Alpert Medical School of Brown University, Providence, RI, USA

**Agnieszka Slowik, MD, PhD**
Department of Neurology,

Jagiellonian University Medical College, Krakow, Poland

**Jennifer A. Smith, PhD, MPH**

Department of Epidemiology, School of Public Health, University of Michigan, Ann Arbor, MI, USA

**Martin Söderholm, MD**

Lund University, Department of Clinical Sciences, Malmö University Hospital, Malmö, Sweden

**Carolina Soriano, Bsc, PhD**

Department of Neurology, Neurovascular Research Group (NEUVAS)

IMIM-Hospital del Mar (Institut Hospital del Mar d’Investigacions Mèdiques), Barcelona, Spain

**Mary J. Sparks, RN, BSN**

Department of Neurology, University of Maryland School of Medicine, Baltimore, MD, USA

**Tara Stanne, PhD**

Institute of Biomedicine, the Sahlgrenska Academy at University of Gothenburg, Gothenburg, Sweden

**Kari Stefansson, MD, PhD**

deCODE Genetics/Amgen, Reykjavik, Iceland

Faculty of Medicine, University of Iceland, Reykjavik, Iceland

**O. Colin Stine, PhD**

Department of Epidemiology and Public Health, University of Maryland School of Medicine, Baltimore, MD

**Konstantin Strauch, PhD**

Institute of Genetic Epidemiology, Helmholtz Zentrum München - German Research Center for Environmental Health, Neuherberg, Germany

Institute of Medical Informatics, Biometry and Epidemiology, Chair of Genetic Epidemiology, Ludwig-Maximilians-Universität, Munich, Germany

**Jonathan Sturm, MBChB, PhD, FRACP**

Department of Neurosciences, Gosford Hospital, NSW, Australia

**Cathie LM Sudlow, DPhil, FRCP(E)**

Centre for Clinical Brain Sciences & Institute of Genomic and Molecular Medicine, University of Edinburgh, UK

**Salman M. Tajuddin, MD, PhD, MP**H

Laboratory of Epidemiology and Population Science, National Institute on Aging, National Institutes of Health, Baltimore, MD, USA

**Robert L. Talbert, PharmD**

College of Pharmacy, University of Texas at Austin, Austin, Texas, USA

**Turgut Tatlisumak, MD, PhD**

Institute of Neuroscience and Physiology, Sahlgrenska Academy at University of Gothenburg, Gothenburg, Sweden

Department of Neurology, Sahlgrenska University Hospital, Gothenburg, Sweden Department of Neurology, Helsinki University Central Hospital, Helsinki, Finland

**Vincent Thijs, MD, PhD**

KU Leuven - University of Leuven, Department of Neurosciences, Experimental Neurology and Leuven Research Institute for Neuroscience and Disease (LIND), Leuven, Belgium

VIB, Vesalius Research Center, Laboratory of Neurobiology, Leuven, Belgium University Hospitals Leuven, Department of Neurology, Leuven, Belgium Gudmar Thorleifsson, PhD

deCODE Genetics/Amgen, Reykjavik, Iceland

**Unnur Thorsteindottir, PhD**

deCODE Genetics/Amgen, Reykjavik, Iceland

Faculty of Medicine, University of Iceland, Reykjavik, Iceland

**Steffen Tiedt**

Institute for Stroke and Dementia Research, Klinikum der Universität München, Ludwig-Maximilians University, Munich, Germany

**Matthew Traylor, PhD**

Department of Clinical Neurosciences, University of Cambridge, Cambridge, UK

**Stella Trompet, PhD**

Department of Cardiology, Leiden University Medical Center, Leiden, The Netherlands

Department of Gerontology and Geriatrics, Leiden University Medical Center, Leiden, The Netherlands

**Valerie Valant, BA**

University of Massachusetts Medical School, Worcester, MA, USA

**Melanie Waldenberger, PhD**

Research unit of Molecular Epidemiology, Helmholtz Zentrum München - German Research Center for Environmental Health, Neuherberg, Germany

Institute of Epidemiology II, Helmholtz Zentrum München - German Research Center for Environmental Health, Neuherberg, Germany

DZHK (German Centre for Cardiovascular Research), partner site Munich Heart Alliance, Munich, Germany

**Matthew Walters, MD**

Institute of Cardiovascular and Medical Sciences, University of Glasgow, UK

**Liyong Wang, PhD**

John P. Hussman Institute for Human Genomics, Miller School of Medicine, University of Miami, Miami, FL, USA

**Xin-Qun Wang, MS**

Department of Public Health Sciences, University of Virginia, Charlottesville, VA

**Sylvia Wassertheil-Smoller, PhD**

Department of Epidemiology and Population Health, Albert Einstein College of Medicine, Bronx, NY, USA

**David R. Weir, PhD**

Survey Research Center, University of Michigan, Ann Arbor, MI, USA

**Kerri L. Wiggins, MS, RD Cardiovascular Health Research Unit**

Department of Medicine, University of Washington, Seattle, WA, USA

**Stephen R. Williams PhD**

Center for Public Health Genomics, Univeristy of Virginia, Charlottesville, VA, USA

**Dorota Wloch-Kopec MD, PhD**

Department of Neurology, Jagiellonian University Medical College, Krakow, Poland

**Daniel Woo, MD, MS**

University of Cincinnati College of Medicine, Cincinnati, OH, USA

**Rebecca Woodfield, MBBChir, MRCP(UK)**

Centre for Clinical Brain Sciences, University of Edinburgh, UK

**Ona Wu, PhD**

Athinoula A. Martinos Center for Biomedical Imaging, Department of Radiology, Massachusetts General Hospital, Charlestown, MA, USA

Department of Radiology, Harvard Medical School, Boston, MA, USA

**Huichun Xu, MD, PhD**

Department of Medicine, University of Maryland School of Medicine, Baltimore, MD, USA

**Alan B. Zonderman, PhD**

Laboratory of Personality and Cognition, National Institute on Aging, National Institutes of Health, Baltimore, MD, USA

**Bradford B. Worrall, MD, MSc**

Departments of Neurology and Public Health Sciences, University of Virginia, Charlottesville, VA, USA

**Paul I.W. de Bakker, PhD**

Department of Medical Genetics, University Medical Center Utrecht, Utrecht, The Netherlands

Department of Epidemiology, University Medical Center Utrecht, Utrecht, The Netherlands

**Steven J. Kittner, MD, MPH**

Department of Neurology, University of Maryland School of Medicine and Veterans Affairs Maryland Health Care System, Baltimore, MD, USA

**Braxton D. Mitchell, PhD, MPH**

Division of Endocrinology, Diabetes and Nutrition, University of Maryland School of Medicine, Baltimore, MD, USA

Geriatric Research and Education Clinical Center, Veterans Administration Medical Center, Baltimore, MD, USA

**Jonathan Rosand, MD, MSc**

Program in Medical and Population Genetics, Broad Institute of MIT and Harvard, Cambridge MA, USA

Department of Neurology and Center for Human Genetic Research, Massachusetts General Hospital, Boston, MA, USA

Department of Neurology, Harvard Medical School, Boston, MA, USA.

Short list of WHI Investigators
**Program Office:** (National Heart, Lung, and Blood Institute, Bethesda, Maryland) Jacques Rossouw, Shari Ludlam, Joan McGowan, Leslie Ford, and Nancy Geller.

**Clinical Coordinating Center:** (Fred Hutchinson Cancer Research Center, Seattle, WA) Garnet Anderson, Ross Prentice, Andrea LaCroix, and Charles Kooperberg.

**Investigators and Academic Centers:** (Brigham and Women's Hospital, Harvard Medical School, Boston, MA) JoAnn E. Manson; (MedStar Health Research Institute/Howard University, Washington, DC) Barbara V. Howard; (Stanford Prevention Research Center, Stanford, CA) Marcia L. Stefanick; (The Ohio State University, Columbus, OH) Rebecca Jackson; (University of Arizona, Tucson/Phoenix, AZ) Cynthia A. Thomson; (University at Buffalo, Buffalo, NY) Jean Wactawski-Wende; (University of Florida, Gainesville/Jacksonville, FL) Marian Limacher; (University of Iowa, Iowa City/Davenport, IA) Jennifer Robinson; (University of Pittsburgh, Pittsburgh, PA) Lewis Kuller; (Wake Forest University School of Medicine, Winston-Salem, NC) Sally Shumaker; (University of Nevada, Reno, NV) Robert Brunner.

**Women’s Health Initiative Memory Study:** (Wake Forest University School of Medicine, Winston-Salem, NC) Mark Espeland.
